# Bi-allelic *ATG12* variants impair autophagy and cause a neurodevelopmental disorder

**DOI:** 10.1101/2025.10.30.25338537

**Authors:** James Lambton, Shotaro Asano, Yuxiang Huang, Fumi Suomi, Tomoya Eguchi, Magali Prigent, Cassidy Petree, Kevin Huang, Aliza Imam, Thomas J. McCorvie, Daniel Warren, Emma Hobson, Helen McCullagh, Doriana Misceo, Anna Bjerre, Marie F. Smeland, Claus Klingenberg, Eirik Frengen, Swati Naik, Gavin Ryan, Annapurna Sudarsanam, Katherine Foster, Pradeep Vasudevan, Rajib Samanta, Fatima Rahman, Shazia Maqbool, Vrajesh Udani, Henry Houlden, Robert McFarland, Jack J. Collier, Reza Maroofian, Wyatt W. Yue, Gaurav K. Varshney, Daniel J. Klionsky, Renaud Legouis, Thomas G. McWilliams, Noboru Mizushima, Monika Oláhová, Charlotte L. Alston, Robert W. Taylor

**Author notes:** **Correspondence to:** Professor Robert W. Taylor **Full address:** Mitochondrial Research Group, Translational and Clinical Research Institute, Faculty of Medical Sciences, Newcastle University, Newcastle upon Tyne, NE2 4HH **E-mail:**.

## Abstract

Macroautophagy/autophagy is an essential developmental and homeostatic process, defined by the endolysosomal degradation of intracellular components and pathogens. Dysfunctional autophagy is implicated in complex human disease, yet reports of congenital autophagy disorders were considered exceedingly rare until the recent report of several unrelated families with bi-allelic variants in *ATG7*, encoding a core autophagy effector complementing the report of two individuals harbouring *ATG5* variants. We now report six affected individuals from five families with bi-allelic *ATG12* variants with complex neurological phenotypes overlapping those of *ATG5* and *ATG7* patients, including developmental delay, congenital ataxia and cerebellar hypoplasia. Structural modelling implicated a potential disruption of the important ATG12–ATG5-ATG16N-ATG3 complex. Biochemical analyses of patient-derived primary fibroblasts from members of two affected families confirmed the loss of stable ATG12–ATG5 conjugate in one family and altered autophagic flux in one unrelated family. HeLa cell models demonstrate a decrease in ATG12–ATG5 conjugate and reduced autophagic flux in response to starvation using the Halo Tag processing assay. Yeast complementation studies demonstrated that equivalent missense *atg12* variants were unable to fully recover the biochemical defect in *atg12* null yeast and also demonstrated a reduced delivery of autophagy substrates to the yeast’s degradative compartment. Functional studies in zebrafish models demonstrated global developmental delay, impaired brain function, and pre-adulthood lethality. Our findings underscore the pivotal role of efficient autophagy in maintaining human neural integrity and emphasise the importance of this emerging group of congenital autophagy disorders, thereby expanding our understanding of adaptive homeostasis in human health and disease.

## Introduction

Autophagy is a cellular degradation and recycling process that sustains eukaryotic homeostasis. During autophagy, cellular cargo is captured within double membrane-bound vesicles known as autophagosomes, which fuse with acidic endolysosomes to form degradative autolysosomes.^1^ At the molecular level, a highly conserved set of core autophagy-related (*ATG*) genes encode proteins that govern phagophore biogenesis, their maturation to autophagosomes and their subsequent degradation by endolysosomes or vacuole. Briefly, upstream triggers are sensed and integrated by the unc-51 like autophagy activating kinase 1 (ULK1) complex initiating phagophore formation *via* the PIK3C3/VPS34-containing class III phosphatidylinositol 3-kinase.^2^ The E1-like enzymatic activity of homodimeric ATG7 aides the expansion of phagophore membranes *via* two ubiquitin-like conjugation systems. One of these systems involves ATG12, a ubiquitin-like protein that functions within a conjugation cascade to promote autophagosome formation.^3^ ATG7-mediated adenylation of ATG12 results in its transfer to ATG5 *via* the E2-like ATG10, generating the ATG12–ATG5 conjugate at the phagophore membrane. ATG12–ATG5 forms a complex with ATG16L1 that facilitates mammalian Atg8-family protein (ATG8) lipidation, a covalent linkage with phosphatidylethanolamine, allowing insertion into the phagophore membranes and subsequent autophagy.^4,5^

Extensive research over the last two decades has unveiled impairments in different aspects of autophagy across a spectrum of human diseases, spanning from various cancers, neurodegenerative conditions and kidney disorders.^6,7^ Previously, we employed gene agnostic whole exome sequencing (WES) strategies that identified multiple unrelated families with bi-allelic, pathogenic variants in *ATG7*, encoding a core-autophagy effector.^8^ Additionally, a pathogenic homozygous missense variant was previously reported in the *ATG5* gene in two siblings.^9,10^ Cerebellar ataxia, cognitive impairment and delayed motor milestones were consistently observed, emphasising the importance of autophagy for human neural integrity.^8–10^ This is consistent with pre-clinical and developmental studies, where conditional ablation of *Atg5* or *Atg7* in the mouse nervous system leads to neuropathology.^11,12^ Whole-body mutants of core *Atg* genes, including *Atg12*, results in highly penetrant perinatal lethality in mice which might suggest that biallelic loss-of-function *ATG12* variants would be incompatible with human life.^13–15^

Here we report the identification of six affected individuals from five unrelated families with bi-allelic variants in *ATG12*. These children presented with neurodevelopmental and neurological deficits, including hypoplasia of the cerebellar vermis, infantile hypotonia, ataxia, seizures, developmental delay and/or intellectual disability. Both *in silico* and experimental analyses using patient primary fibroblast cells, HeLa cell models and yeast illustrate the deleterious impact of these novel variants, with zebrafish models highlighting their developmental impact, combining to underscore the critical importance of autophagy in human neural development and disease.

## Materials and methods

### Patients

#### Ethics and Consent

Patients and their families have consented to reporting of their data according to the Declaration of Helsinki. Ethical approval was obtained from all centres: Family 1 was identified as part of a National Institute of Health Research (NIHR) funded trio whole exome sequencing project (‘Diagnosis of Mitochondrial Disease without Muscle”; IRAS 255610; REC: 20/NE/0272); Family 2 was identified through a study approved by the Regional Committee for Medical Research Ethics – South-East Norway, REK 2010/1152a; the investigation of Family 5 is covered by UCL IRAS Project-ID:310045. The families and associated researchers were connected through the GeneMatcher platform.^16^

#### Family 1

S1, a female infant, was the first child born to non-consanguineous parents following *in vitro* fertilization (IVF) for unexplained infertility. Born at term, by normal vaginal delivery following an uneventful pregnancy, with their weight in the 25^th^-50^th^ centiles. Newborn examination was normal with the exception of overlapping 4^th^ and 5^th^ toes bilaterally. Feeding difficulties, excessive weight loss and jaundice were noted in early life. Examination revealed hypotonia and apnoea when feeding. Supplemental oxygen and nasogastric tube feeding were commenced, and a subsequent swallowing assessment confirmed a poorly coordinated swallow.

Axial hypotonia with excessive head lag remained a prominent feature on neurological review in early life when myopathic facies and a head circumference in the 9-25^th^ centiles were also recorded. Limb movements were markedly decreased, and she was unable to fix and follow. Cranial magnetic resonance imaging (MRI) performed 6 months apart revealed diffuse parenchymal volume loss involving both grey and white matter (**Fig**. **1A**). The corpus callosum was hypoplastic with marked volume loss of cerebellar hemispheres and vermis between MRIs. The mamillary bodies were absent and the hippocampi simplified with delayed myelination. S1 died in early life due to her rapidly progressive neurodegenerative condition with refractory epilepsy.

**Figure 1.**
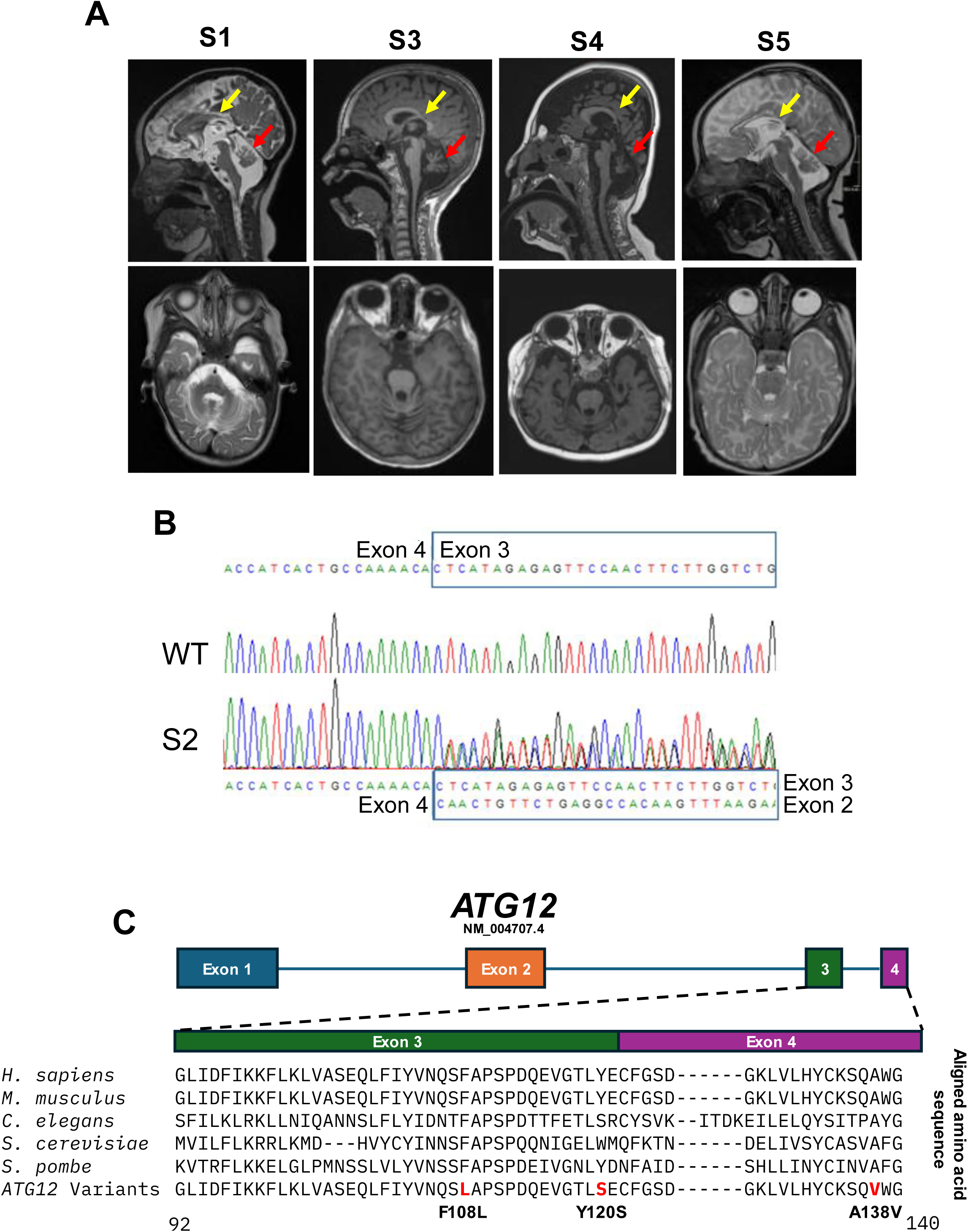
Brain magnetic resonance imaging and identification of *ATG12* variants. (**A**) Brain MRIs of S1 in the first year of life (T2 weighted), S3 (T1 weighted), S4 (T1 weighted) and S5 (T2 weighted) showing posterior atrophy of the corpus callosum and cerebellar vermian hypoplasia, denoted by yellow and red arrows, respectively. (**B**) Sanger sequencing of S2 cDNA showing splicing defects due to the c.363+3A>T variant. The double peaks detected are in line with the sequences of *ATG12* exon 2 and 3 as indicated (**C**) Schematic of ATG12 (RefSeq accession number NM_004707.4), with multiple sequence alignment of orthologues from various species, aligned using Clustal Omega, demonstrating the conservation of the amino acid residues involved.

Further clinical review by paediatric neurology led to a range of neurological, metabolic, genetic and radiological investigations that were diagnostically uninformative. Blood and fibroblasts were taken for cytogenetic (array CGH) and molecular genetic analysis.

S2, a male infant and younger sibling of S1, was born by normal vaginal delivery with weight in the 50^th^–75^th^ centiles with a head circumference in the 25^th^-50^th^ centiles. No resuscitation was required at birth, but hypotonia and paucity of movement was noted. He had bilateral positional talipes with overlapping 4^th^ and 5^th^ toes on the right foot. Tachypnoea with low oxygen saturation prompted admission to the neonatal intensive care unit where he was initially treated for transient tachypnoea of the newborn. A persistent oxygen requirement and feeding difficulties prompted further medical investigations.

Recurrent seizures were treated with levetiracetam in the first months of life. A cranial MRI at soon after birth demonstrated a structural malformation with deep sulcation of the right occipital lobe and generalised (particularly vermian) cerebellar and corpus callosal hypoplasia. A comparison of S1 and S2 scans suggested a number of common features, most notably the vermian volume loss and callosal hypoplasia.

Ongoing feeding difficulties prompted insertion of a gastrostomy in their first year of life. Around this time increased seizure activity was noted and while initial EEGs had been unremarkable, by 4 months the EEG was consistent with epileptic encephalopathy. Blood and fibroblasts were taken for cytogenetic (array CGH) and molecular genetic analysis. S2 died in the first year of life following a presumed viral infection and subsequent increase in apnoeic episodes. The cause of death was recorded as a neurodegenerative condition with refractory epilepsy as in the case of S1, the older sibling.

#### Family 2

S3, a male, is the fourth child of healthy consanguineous parents. His two oldest siblings are healthy. One ol had difficulty with walking and speaking, impaired vision with nystagmus and died from kidney problems in childhood. Many of these features were similar to S3, suggesting she may have had the same disease although no formal genetic studies were performed. His mother also reported previous pregnancy losses.

Pregnancy, term delivery and the neonatal period of S3 were normal. Limited detail is available on his early life, but psychomotor delay became evident during the first year and despite experiencing a single seizure he did not go on to develop epilepsy.

He was diagnosed with kidney disease, anaemia and thrombocytopenia. He received peritoneal dialysis for five years until a renal transplant was performed. Renal ultrasound and renal MRI before the transplant showed bilateral atrophic kidneys with small cysts.

He was severely growth retarded with Z-score for length at -6 SD before the renal transplant. Growth remained impaired and did not improve substantially after transplantation. He has been diagnosed with hypothyroidism and receives thyroxine replacement therapy.

He has limited verbal language skills, moderate intellectual disability and wears hearing aids to assist with bilateral sensorineural hearing impairment. He walks only with the aid of a walker due to ataxia. Brain MRI obtained in childhood showed atrophy of both cerebellar hemispheres and the vermis and widespread bilateral polymicrogyria (**Fig**. **1A**). An ophthalmological examination revealed bilateral optic atrophy and typical Bull’s eye maculopathy.

His clinical appearance in adolescence is one of short stature, with a slender build. He has microcephaly and mild dysmorphic features including a broad nasal bridge, anteverted nostrils, long philtrum and cupid’s bow upper lip. His ears are low set and he has sparse scalp hair. He also has bilateral fifth finger clinodactyly of both hands.

#### Family 3

S4 is the first child of consanguineous parents (**Fig. 1A**). He was born by lower segment caesarean section following a pregnancy complicated by gestational diabetes mellitus. An antenatal diagnosis of dilated urinary bladder was not confirmed on postnatal abdominal ultrasound scan, which was reported normal. Parents believed that he was well until concerns were raised at a six-week check by the health visitor who felt that he was not fixing and following. In early life he experienced his first seizure, which lasted for more than 45 minutes. Currently, he is on levetiracetam, vigabatrin and gabapentin along with the ketogenic diet.

#### Family 4

S5 is a male child of healthy non-consanguineous parents. A sibling died of atypical brainstem glioma in childhood. Pregnancy with S5 followed a natural conception and proceeded without incident until a fetal ultrasound revealed talipes and dilated ventricles. After birth he was noted to have neonatal jaundice and poor feeding. He received phototherapy and was discharged home. He was readmitted soon afterwards with abnormal movements. Although EEG was reported normal, he went on to have clinical seizures and brain MRI scan revealed dysgenesis of the corpus callosum, prominent cavum septum pellucidum, abnormal cerebellar vermis, small cerebellar hemispheres and a small posterior fossa (**Fig. 1A**). In view of the above features, microarray analysis was requested which did not find any copy number variation. Rapid WES was then requested under R14 category as per the NHS England National Genomic Test Directory and the result was normal. Metabolic investigations including blood lactate, organic acids, amino acids, carnitine, very long chain fatty acids were normal. Further genomic panel testing was undertaken, with normal results.

Clinical review in childhood showed significant hypotonia, profound developmental delay with a non-verbal and non-mobile child who has a gastrostomy *in situ*, intractable epilepsy and a movement disorder. Palliative care has been involved.

#### Family 5

S6 is male and the third child of consanguineous parents. He presented in early life with concerns regarding developmental delay, failure to thrive and seizures soon after birth. Seizures were controlled with levetiracetam. Antenatal and post-natal history were not significant. He was born at term via spontaneous vaginal delivery. He was partially vaccinated and had multiple hospital admissions with uncontrolled seizures. An older sibling had impaired vision with borderline intellectual disability, while anotherhad a history of urethral meatal stenosis but no developmental delay. A younger sibling had global developmental delay, behavioural issues and recurrent chest and gastrointestinal infections.

Examination in early childhood showed clinical signs of malnutrition but no obvious dysmorphic features. He was floppy with poor truncal control, head lag and hyporeflexia. He appeared unable to grasp objects placed in his palms. He could fix on familiar faces but was unable to follow. Over time, he developed spasticity with failure to gain weight. Speech did not progress beyond simple vocalisations. EEG in early life was normal, but no neuroimaging was obtained. He was emaciated, weighing in the <0.4^th^ centile, and passed away following a period of vomiting leading to severe dehydration.

### Genetic Analysis

#### Whole-Exome Sequencing (WES)

All subjects underwent genetic investigations using EDTA blood-derived DNA. For Family 1 (S1 and S2), gene agnostic WGS of S1 and her parents was performed via the National Health Service (NHS) testing pathway (R14: acutely unwell children with a likely monogenic disorder). Subsequently, S2 and his parents were recruited to a National Health and Care Research (NIHR) funded trio whole exome sequencing project for research-based analysis. Concurrent singleton whole-exome sequencing was performed for S1 to allow comprehensive data analysis to be performed as a quad, with both affected children being analysed alongside both unaffected parents. EDTA blood DNA from S1, S2 and their parents underwent library preparation using the Twist Exome 2.0 reagent (Twist Biosciences) and sequencing on an Illumina NovaSeq 6000 sequencer using 2×150 bp paired-end reads. Reads were aligned to the GRCh38 human genome build using the Burrows Wheeler Aligner (BWA)^17^; variant calling was performed using the GATK HaplotypeCaller (version 3.4). All variants were annotated using the Ensembl Variant Effect Predictor (VEP), and a minor allele frequency of MAF<0.01 (gnomAD v3.1) was used to define the rare variant dataset for each member of the quad for Family 1.^18^ Each variant in S1 and S2 was first denoted as maternal or paternal, then genes with biallelic variants segregating between the two siblings were determined. SpliceAI was used to predict the likely impact of putative splicing variants.

For Family 2 (S3), singleton whole-exome sequencing was performed. FASTQ files were aligned to the GRCh38 human genome reference sequence using Burrows-Wheeler Aligner (BWA) v.0.7.8. with duplicate reads removed using Picard v.1.119 (http://broadinstitute.github.io/picard/), indel realignment, base quality recalibration and variant calling was performed using Genome Analysis Toolkit (GATK) v.4.2.4.1.^17,19,20^ Variant annotation was performed using the Ensembl Variant Effect Predictor (VEP).^18^ The variant calling file (VCF) was analysed using the FILTUS program.^21^ Variants with allelic frequency >0.01 in gnomAD v3.1.2 (gnomad.broadinstitute.org) or with a Combined Annotation Dependent Depletion (CADD) PHRED score <15 *in silico* tool were discarded.^22,23^ Finally, prioritization of variants focused on missense, nonsense, frameshift, and small insertion/deletion variants, with an anticipated autosomal recessive (homozygous and compound heterozygous) and autosomal dominant (*de novo*) mode of inheritance.

#### Sanger sequencing confirmation of *ATG12* variants

Blood-derived genomic DNA was subject to PCR amplification and Sanger sequencing to confirm the presence of the identified *ATG12* variants (**Supplementary Table 1**). PCR products were purified using *exo*I and shrimp alkaline phosphatase (Promega) and Sanger sequenced using the ABI BigDye terminator cycle-sequencing kits v3.1 (Life Technologies, Carlsbad, CA) before capillary electrophoresis using an 3730xl DNA analyser running proprietary DNA Sequencing Analysis Software v. 5.1 (Applied Biosystems, Foster City, CA). Sequences were visualised using FinchTV (Family 1) and SeqScape Software v.2.7 (Family 2) (Thermo Fisher Scientific).

#### RNA studies to investigate a putative splicing variant in *ATG12*

Complimentary DNA (cDNA) was derived from whole RNA extracted from patient and age-matched control fibroblasts cultured in standard Dulbecco’s modified Eagle’s medium (DMEM). Reverse transcription was performed using Moloney Murine Leukemia Virus reverse transcriptase reverse transcription system and random hexamer primers (Promega). Primers designed to amplify cDNA spanning from exon 1 of *ATG12* through to its 3’-UTR were used for PCR amplification (**Supplementary Table 2**). Electrophoretic separation of the PCR amplicons using a 2% agarose gel facilitated identification of normal- and abnormal-length cDNA fragments. Amplicons were subject to DNA sequencing to determine the effect of the *ATG12* variant (c.363+3A>T) on mRNA splicing.

### Protein Modelling

The previously described crystal structure for the ATG12–ATG5-ATG16N-ATG3 complex was obtained from RCSB PDB (4NAW).^24^ To generate a model of the full length ATG12– ATG5-ATG16N-ATG3 complex, the canonical full-length sequences were obtained from UniProtKB with the accession numbers Q9H1Y0-1, O94817-1, Q9NT62-1 and Q676U5-1, respectively.^25^ The multimeric complex was then modelled using AlphaFold-MULTIMER using standard settings.^26^

### Assessment of autophagy in Subject-derived cell lines

#### Immunoblotting analysis of subject fibroblasts

Primary subject fibroblasts were cultured in Gibco^®^ DMEM (4.5 g/L glucose, 2 mM L-glutamine, 1 mM pyruvate) with 10% fetal bovine serum (FBS), 50 μg/mL uridine, 1x non-essential amino acids, 1x penicillin-streptomycin and incubated at 37°C in 5% CO_2_ and lysed using cold lysis buffer (50 mM Tris/HCl pH 7.4, 130 mM NaCl, 2 mM MgCl_2_, 1 mM PMSF, 1% Nonidet P-40, one cOmplete^TM^ EDTA-free protease inhibitor tablet [Roche]). For autophagy flux assays, autophagy was induced by addition of 1 μM AZD8055 with late-stage autophagy being blocked with 100 μM chloroquine. SDS-PAGE was performed using 8% or 12% polyacrylamide gels.

Membranes were blocked for one hour at room temperature in 5% skimmed milk power (Marvel) dissolved in tris buffered saline (Santa Cruz) with 0.1% Tween-20 (Sigma) (TBS-T). Immunoblotting was performed either overnight at 4°C or for one hour at room temperature with primary antibodies. Immunoblotting was performed using the following antibodies: ATG12 1:1000 (Abcam ab303488), ATG7 1:1000 (Cell Signalling Technology #8558), ATG3 1:500 (Cell Signalling Technology #3414), ATG5 1:500 (Cell Signalling Technology #2630), SQSTM1/p62 1:1000 (Abcam ab109012), MAP1LC3B/LC3B 1:1000 (Cell Signalling Technology #2775), GAPDH 1:10000 (Proteintech 60004-1-Ig), anti-rabbit 1:2000 (DAKO P0399), anti-mouse 1:2000 (DAKO P0260). Following this, membranes were incubated in species specific HRP-conjugated secondary antibody for 1 hour at room temperature. Membranes were visualised using SuperSignal West Pico PLUS Chemiluminescent Substrate (Thermo Scientific) and imaged with a ChemiDoc XRS+ Imaging system using Image Lab Software (BioRad).

#### Patient fibroblast autophagy flux assay

Primary patient and age-matched control fibroblasts were seeded at 100,000 cells per 60-mm dish and cultured for 2 to 3 days as described to reach ∼ 60% confluency. Subsequently, cells were treated with an unsaturating (2 nM) or saturating (100 nM) concentration of bafilomycin A_1_ (BafA_1_; Enzo Life Sciences) alongside basal conditions or starvation (Hanks’ balanced salt solution [HBSS; Gibco]) conditions and 37°C, 5% CO_2_. Cells were harvested after 3 h for immunoblot analysis.

#### Patient fibroblast bulk autophagic sequestration activity assay (LDH sequestration assay)

Autophagic sequestration assays were performed by measuring the activity of autophagosomal lactate dehydrogenase (LDH) as previously described^27,28^. Subject cells were cultured using standard tissue culture conditions, at 37°C, 5% CO_2_ in DMEM with high glucose (Gibco^®^), supplemented with 1X GlutaMAX, 10% FBS and 100 U/ml penicillin-streptomycin. To induce autophagy, cultures were acutely deprived of serum and amino acids using EBSS with 100 nM BafA_1_ for 3 h at 37°C, 5% CO_2_. Cells were harvested using 0.25% trypsin-EDTA and collected in PBS containing 5% BSA. After centrifugation at 500 x *g* for 5 min at 4°C, cells were resuspended in sucrose solution (10% w:v, ice cold). Subsequently, plasma membrane disruption was accomplished using electroporation (Bio-Rad, 2kV, 25 µF, and 400 Ω with a pulse of ∼8 ms duration). Cells were resuspended in 400 µl phosphate-buffered sucrose (100 mM sodium monophosphate, 2 mM EDTA, 2 mM DTT, 1.75% sucrose, pH 7.5). Following this, 550 µl of the disrupted cell suspension was resuspended in 900 µl ice-cold resuspension buffer (50 mM sodium monophosphate, 1 mM EDTA, 1 mM DTT, pH 7.5) for the sedimented measurement (LDH_‘Sediment’_). Autophagic vacuoles were then sedimented by centrifugation at 18,000 x *g* for 45 min at 4°C. The resulting supernatant was aspirated and flash frozen using liquid nitrogen and stored at -80°C.

For total LDH measurement, 100 µl from the disrupted cell suspension solution was collected, flash frozen with liquid nitrogen and stored at -80°C (LDH_‘Total’_). LDH_”Sediment”_ was diluted in resuspension buffer with 1% Triton X-405 and LD_‘Total’_ with 1.5% Triton X-405 in a cold room for 30 min with agitation. Subsequently, samples were centrifuged at 18,000 x *g* for 5 min at 4°C, after which LDH activity was measured as described previously in a working solution containing 0.6 mM pyruvate and 0.36 mM NADH.^27,28^

### Assessment of autophagy in HeLa *ATG12* knockout models

#### Cell culture

HeLa cells, which were authenticated by RIKEN, were used in this study. Cells were cultured in DMEM (Sigma-Aldrich) supplemented with 10% FBS (Sigma-Aldrich) at 37°C, 5% CO_2_. For starvation, cells were treated with amino acid-free DMEM without FBS.

#### Reagents and antibodies

Anti-HA (Cell Signaling Technology #3724), anti-ATG12 (MLB M154-3), anti-HSP90 (BD Transduction Laboratories610419), anti-LC3A/B (Cell Signaling Technology #12741), and anti-SQSTM1/p62 (PROGEN GP62-C) antibodies were used as primary antibodies. HRP-conjugated anti-mouse IgG (Jackson ImmunoResearch Laboratories 111-035-003), HRP-conjugated anti-rabbit IgG (Jackson ImmunoResearch Laboratories 111-035-144), and HRP-conjugated anti-guinea pig IgG (Jackson ImmunoResearch Laboratories 106-035-003) antibodies were used as secondary antibodies.

#### Generation of *ATG12* knockout HeLa cells

An *ATG12* knockout HeLa cell line was generated using the CRISPR-Cas9 gene editing technique. A CRISPR guide RNA (gRNA) targeting *ATG12* (exon 1: 5’-CTTCCTACTTCAATTGCTGC-3’) was inserted into pSpCas9(BB)-2A-Puro (PX459) V2.0 plasmid (a gift from Feng Zhang; Addgene plasmid #62988; http://n2t.net/addgene:62988; RRID:Addgene_62988) and transfected into HeLa cells using Lipofectamine 2000 (Thermo Fisher Scientific) following the manufacturer’s protocol.^29^ Puromycin (Sigma) selection was started 1.5 days after transfection. Knockout clones were obtained by cell sorting using CytoFLEX SRT (Beckman Coulter Life Science).

#### Constructs and plasmids

The human *ATG12* cDNA (NM_004707.4), inserted with intron 3, was cloned into pMRX-IPU with an N-terminal 3×HA tag. *ATG12* variants were generated by site-directed mutagenesis. *LC3* was inserted into pMRX-IBU-HaloTag7 to create pMRX-IBU-HaloTag7-LC3.

#### Retrovirus infection

HEK293T cells were transfected with retroviral plasmids together with packaging and envelope plasmids (pCG-gag-pol and pCG-VSVg) using Lipofectamine 2000 (Thermo Fisher Scientific), according to the manufacturer’s protocol. Three days after the transfection, the retrovirus-containing medium was collected with a 0.45-µm pore filter and transferred to the recipient cells. After two days, selection was performed with 2 µg/mL puromycin (Sigma), 2.5 µg/mL blasticidin (Fujifilm Wako Pure Chemical Corporation), or 50 µg/mL hygromycin (Nacalai Tesque).

#### HaloTag processing assay

Cells were pulse-labelled with tetramethylrhodamine-conjugated ligand (100 nM) for 20 min, and the ligands were washed out. After starvation, cells were lysed in a lysis buffer (PBS containing 1% Triton X-100 and protease inhibitors; Nacalai Tesque), and incubated on ice for 30 min. The lysates were collected and centrifuged at 12,000 × *g* for 15 min. The supernatant was collected and separated by SDS-PAGE. The gel was analysed with Odyssey (LI-COR) immediately after SDS-PAGE.^30^

#### Immunoblotting

Cell lysates were separated by SDS-PAGE and transferred onto polyvinylidene fluoride (PVDF) membranes. The membranes were blocked with 4% non-fat milk powder in Tris-buffered saline with 0.1% Tween 20 and incubated with primary antibodies at 4°C overnight. After washing, the membranes were incubated with species-appropriate secondary antibodies (1:10,000) at room temperature for 1 h. Signals were detected using FUSION Solo.7S.EDGE (Vilber Lourmat) with Immobilon Western Chemiluminescent HRP Substrate (Millipore).

### Assessment of autophagy in *Saccharomyces cerevisiae Atg12* mutants

#### Yeast strains, media, and growth conditions

Yeast strains used in this study are listed in **Supplementary Table 1**. Yeast cells were cultured at 30°C in rich medium (YPD; 1% yeast extract, 2% peptone, and 2% glucose). To induce autophagy, cells in the mid-log phase (OD_600_ = 0.8-1.0) were shifted to nitrogen-starvation medium with glucose (SD-N; 0.17% yeast nitrogen base without ammonium sulfate or amino acids, and 2% glucose) for the indicated times.

#### Autophagic flux assays and western blotting

Atg8–PE lipidation, GFP-Atg8 processing, Pho8Δ60 activity, and Pgk1-GFP processing assays were performed as previously described.^31–33^ Antisera were from the following sources: Atg8,^34^ Pgk1 (a generous gift from Dr. Jeremy Thorner, University of California, Berkeley), monoclonal YFP (Clontech, 632381), monoclonal HA (Sigma, H3663). The blot was imaged using a ChemiDoc Touch imaging system (Bio-Rad) and quantified using Bio-Rad Image Lab software.

#### Yeast strains and growth media for fluorescence microscopy

*Saccharomyces cerevisiae* strains used in this study are listed in **Supplementary Table 3**. OC588 (BY4742, *atg8*::*GFP-ATG8-URA3*, *ura3Δ0, his3Δ1, leu2Δ0, lys2Δ0*) was obtained by integration of plasmid *pP_1K_GFP-ATG8(406)* at the BY4742 chromosomal *ATG8* locus.^33^ The *GFP-ATG8 atg12*Δ (OC744) strain was created by genomic integration of a deletion cassette PCR fragment amplified from the OC751 strain in the *GFP-ATG8* (OC588) strain.

Yeast cells were grown to log phase in complete synthetic medium (CSM: 0.17% yeast nitrogen base, 0.5% ammonium sulfate, 2% glucose and amino acids without uracil or leucine). YNB-N (0.17% yeast nitrogen base without ammonium sulfate and amino acids and 2% glucose) was used as nitrogen starvation medium.

The *ATG12* gene with 500 bp upstream and downstream regions was amplified by PCR, cloned into the TA pCR™4-TOPO® vector (ThermoFisher Scientific) and sequence verified. Plasmids encoding Atg12^W166S^ or Atg12^A184V^ were generated using the Phusion™ Site-Directed Mutagenesis kit (ThermoScientific) in pCR4-*ATG12*. Wild-type (WT) *ATG12* and mutant sequences were cloned into the pRS315 vector at *Xba*I and *Pst*I sites (**Supplementary Table 4**).

#### Fluorescence microscopy

Cells were grown to log phase in CSM without uracil and leucine and then were transferred to nitrogen-starvation medium for 4 h. At different time points, cells were collected and observed with a three-dimensional deconvolution microscope DMIRE2 (Leica Microsystems) equipped with an HCxPL APO 100 × oil CS objective, NA = 1.40 (Leica Microsystems) and an incubation chamber. Images were captured with a 20-MHz Cool SNAPHQ2 charge-coupled device camera (Roper Technologies), with a z-optical spacing of 0.2 μm. Metamorph software (Molecular Devices) was used to acquire z-series and to deconvolve the images.

#### Statistical analysis

For the bulk autophagic sequestration assay, ordinary one-way analysis of variance and Sidak’s multiple comparison tests were performed to assess the differences between samples. Graphs and statistics for yeast data were performed using GraphPad Prism 8.0 software. The tests used Kruskal-Wallis or two-sided Fisher’s exact test, and the statistical significance is indicated in the figure legends.

### Developmental assessment of *atg12* null zebrafish

#### Ethics Statement and Zebrafish Husbandry

All procedures involving experimental animals were conducted in accordance with institutional policies and NIH guidelines. Zebrafish (*Danio rerio*) were housed and maintained under standard conditions in an AAALAC-accredited facility at the Oklahoma Medical Research Foundation (OMRF). All experimental protocols were reviewed and approved by the OMRF Institutional Animal Care and Use Committee (IACUC; protocol #22-76).

#### Generation of *atg12* knockout zebrafish

We used the CRISPR/Cas9 method to generate *Atg12* knockout animals according to established protocols.^35,36^ Briefly, a guide sequence was designed using the CRISPOR tool, and guide RNA (gRNA) was chemically synthesized by Synthego Inc. (CA, USA). A 6 μL mixture containing 1 μL of 40 μM Cas9-NLS protein (UC Berkeley QB3 Macrolab, Berkeley, CA, USA), 500 ng of gRNA (in 3 μL), and 2 μL of 1 M potassium chloride was injected into one-cell stage embryos.^37^ Founder embryos were raised to adulthood and outcrossed with WT to obtain heterozygous carriers, which were identified by genotyping the F1 progenies using fluorescent PCR.^36^ Heterozygous carriers were subsequently inbred to generate *homozygous* animals for functional analysis, and indels were confirmed by Sanger sequencing.

#### Morphological phenotyping

Phenotyping was performed in homozygous animals by breeding two heterozygous carriers. Progeny from heterozygous crosses were monitored daily until 15 days post-fertilization. The larvae were manually positioned in 2% methylcellulose (Sigma, USA) under a stereomicroscope for visualization and image capture. Bright-field images were captured using a Nikon DS-Fi2 high-definition camera mounted on a Nikon SMZ18 stereomicroscope (Nikon, Japan) equipped with auto-Z-stacking capability. After imaging, each embryo/larva was genotyped as described earlier.^36^

#### Whole-mount immunohistochemistry

Whole-mount immunohistochemistry was performed to label brain anatomical structures following established protocols.^38^ The antibodies used are mouse anti-acetylated-tubulin antibody (1:500, Sigma-Aldrich T7451), rabbit anti-acetylated-tubulin antibody (1:250, Cell Signaling Technology #5335) and mouse anit-SV2A antibody (1:500, DSHB SV2). The secondary antibodies used are goat anti-mouse IgG Alexa Fluor® 647 antibody, goat anti-rabbit IgG Alexa Fluor® 488 antibody (1:500; Jackson ImmunoResearch Laboratories). Samples were mounted in 1.2% agarose and imaged using a Light-sheet microscope.

#### Behavioral assay

All behavioral assays were performed at room temperature (RT) as previously described.^38,39^ For the Light/Dark Transition (LDT) test, 4 days post-fertilization (dpf) larvae were individually placed in 96-well plates containing 150 µL of embryo water. At 5 dpf, the plate was transferred to a Noldus chamber, and locomotor activity was recorded using the DanioVision system with EthoVision XT software (Noldus Information Technology, Leesburg, VA, USA). Following a 30-minute light habituation period, larvae were subjected to two alternating cycles of 30 minutes in darkness and 30 minutes in light. Locomotor activity was quantified as distance traveled (mm/min), and minute-by-minute data were plotted using GraphPad Prism (GraphPad Software, San Diego, CA, USA).

## Results

### Genomic analyses reveal bi-allelic *ATG12* variants

Microarray analysis (array CGH) undertaken for siblings S1 and S2 revealed an 11-kb copy number loss at 9p24.3 (including exons 40-44 of *DOCK8*) for S1 only; there was no phenotypic correlation, so this was not further investigated. Rapid, gene agnostic trio WES analysis for S1 identified bi-allelic variants of unknown significance in *SEMA6B* and *PRMT7*; however, these were not found to segregate with S2. Repeat, research-based WES sequencing was undertaken for S1 and S2 and both parents considering their shared but undiagnosed presentation. Initial investigation of variants that segregated between both siblings and that were biparentally inherited failed to identify any suitable candidates, but by focusing on variants that segregated across the two affected siblings (independent of parental data), two heterozygous variants were identified in *ATG12*. First, *ATG12* (NM_004707.4): c.359A>C; p.(Tyr120Ser) missense variant with a CADD PHRED score of 24.4 and REVEL score of 0.288. The variant has been reported on gnomAD v4.1.0 with 24 heterozygotes recorded, equating to an allele frequency of 2.578x10^-5^. Subsequent analysis of the familial data revealed this variant to be paternally transmitted. Following completion of the functional experiments described in this manuscript, the *ATG12* c.359A>C; p.(Tyr120Ser) variant is now classified as ‘likely pathogenic’ according to the American College of Medical Genetics and Genomics (ACMG) variant classification guidelines, fulfilling the following criteria: PS3_moderate (2 points), PM1_supporting (1 point), PM2_moderate (2 points) and PP4_supporting (1 point).^40,41^ Additionally, an *ATG12* (NM_004707.4): c.363+3A>T putative splice variant was identified that was detected in both children but was not represented in the variant list for either parent. This variant has been reported in gnomAD v4.1.0 with 22 heterozygotes, equating to an allele frequency of 2.373x10^-5^ including one homozygote. The *ATG12* c.363+3A>T putative splice variant is now classified as ‘likely pathogenic’ according to the ACMG variant classification guidelines, fulfilling the following criteria: PM1_supporting (1 point), PM3_moderate (2 points), PM4_moderate (2 points), and PP4_supporting (1 point). The variant has a CADD PHRED score of 24.2 and SpliceAI score of 0.830. Visualisation of the BAM files using the Integrative Genomics Viewer (IGV) allowed visual confirmation of the maternal putative splice variant, and there is an adjacent polynucleotide tract which proffers an explanation as to why the maternal variant was not called. Variants were interpreted according to the ACMG 2015 & 2020 and Association for Clinical Genomic Science (ACGS) 2024 guidelines.^40,41^ The presence of all *ATG12* variants and their segregation across family members was undertaken using Sanger sequencing.

We performed WES of S3, and data analysis according to a recessive mode of inheritance identified the following homozygous missense variant ATG12 (NM_004707.4): c.413C>T; p.(Ala138Val), with a CADD PHRED score of 31 and REVEL score of 0.908. The variant is reported with an allele frequency of 1.12x10-5 in gnomAD v4.1.0, corresponding to 18 heterozygotes. Following completion of the functional experiments described in this manuscript, the ATG12 c.413C>T; p.(Ala138Val) variant is now classified as ‘likely pathogenic’ according to the ACMG variant classification guidelines, fulfilling the following criteria: PM1_supporting (1 point), PM2_moderate (2 points), PM3_supporting (1 point),

PP3_supporting (1 point) and PP4_supporting (1 point).^40,41^ Sanger sequencing of the family trio confirmed the variant to be homozygous in S3 and heterozygous in his healthy parents. No sample was available from a sibling, possibly affected by the same disease. No genetic variants were identified that explained the kidney involvement reported in Family 2, and a separate aetiology is possible.

Having successfully used GeneMatcher to identify one additional patient (S3), the detailed assessment of genomic data within the Genomic Medicine Service (GMS) Clinical Variant Ark (https://ip-cva-help.genomicsengland.co.uk/latest/) – a variant repository from individuals who have undergone WGS studies through the NHS GMS or the 100,000 genomes project (https://www.genomicsengland.co.uk/initiatives/100000-genomes-project) – revealed two additional families: S4 who was homozygous for the c.359A>C, p.Tyr.120Ser *ATG12* variant; and S5 who also harboured the c.359A>C, p.Tyr.120Ser *ATG12* variant *in trans* with a novel c.235C>T, p.Arg79* *ATG12* nonsense variant.

WES of the proband of Family 5 (S6) was performed as previously described.^42^ Following the exclusion of pathogenic or likely pathogenic variants in currently known genes associated with neurological disorders, novel aetiologies were interrogated and a novel homozygous missense variant in *ATG12*, located within a large region of homozygosity, was identified. Segregation analysis confirmed co-segregation of the homozygous c.324T>G, p.Phe108Leu missense variant in *ATG12* with the disease phenotype.

### The c.363+3A>T *ATG12* variant causes aberrant mRNA splicing

Analysis of fibroblast-derived RNA from S2 demonstrated skipping of exon 3 (**Fig**. **1B**); this was predicted to cause an in-frame deletion of exon 3, consisting of 21 residues. The presence of a homozygous individual on gnomAD would suggest a hypomorphic mechanism, in that the c.363+3A>T variant causes ‘leaky’ splicing, and there are sufficient residual wild-type transcripts to compensate. Analysis of the splicing data also supports the mechanism relating to the paternal c.359A>C variant. Given the variant’s proximity to the exon-intron boundary, it was postulated that this could cause aberrant splicing, but this was not supported by SpliceAI (score of 0.03, possible splicing significance is >0.2), or cDNA sequencing data. The c.359A>C variant is therefore likely to cause the predicted tyrosine-to-serine amino acid substitution, p.(Tyr120Ser).

### ATG12 variants may result in disrupted ATG12–ATG5-ATG16N-ATG3 interaction

The three missense variants identified (p.Phe108Leu, p.Tyr120Ser and p.Ala138Val (**Fig. 1C**)) map to the surface of the C-terminal domain of ATG12 which may imply roles in protein interactions (**Fig. 2A**). The crystal structure of ATG12–ATG5, in complex with the N-terminal minimal interacting regions of ATG16L1 (ATG16N) and ATG3, has previously been determined to 2.2 Å resolution (**Fig**. **2B**).^24^ Using AlphaFold-MULTIMER, we expanded the current ATG12–ATG5-ATG16N-ATG3 structure to include full-length ATG3 protein in the modelling.^26,43^ Our expanded model recapitulates the reported interaction surface between ATG12 and ATG5 but reveals beyond the crystal structure how full-length ATG3 could wrap around ATG12 extensively (**Fig**. **2C**).

**Figure 2.**
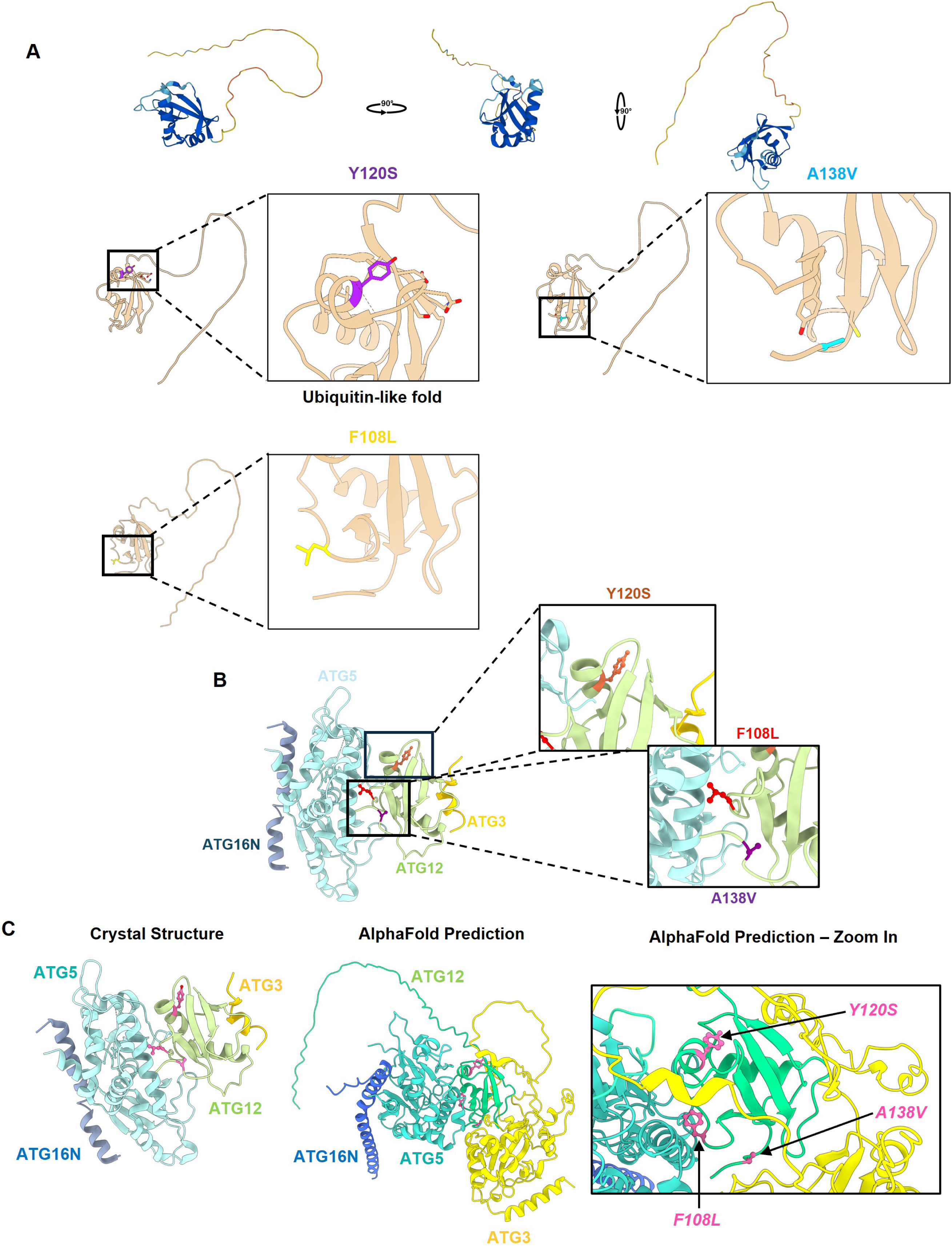
*In silico* structural analysis of ATG12 variants illustrating the effect on complex formation. (**A**) Alphafold prediction of ATG12 structure (Accession number: O98417), in orthogonal views. Insets: Position of p.Tyr120Ser (Y120S) variant in the ubiquitin fold of ATG12 is shown in purple. Position of p.Ala138Val (A138V) variant is shown in blue. Position of the p.Phe108L (F108L) variant is shown in (**B**) Reported crystal structure of the ATG12-ATG5-ATG16N-ATG3 complex (RCSB PDB accession: 4NAW) showing the proximity of the two missense variants in this study to the interface between ATG12 and ATG5. (**C**) Comparison of the existing crystal structure of the ATG12-ATG5-ATG16N-ATG3 complex and our expanded Alphafold model with the two missense variants annotated in pink. The expanded AlphaFold model shows the full length ATG3 sequence (yellow) wrapped around ATG12 (green), placing the variants in close proximity to the interfaces with both ATG5 and ATG3.

The three missense variants identified (p.Phe108Leu, p.Tyr120Ser, and p.Ala138Val) map to different segments of ATG12, all three of which lie close to ATG12’s interfaces with ATG5 and ATG3. The p.Ala138 lies on the external face of ATG12, in very close proximity to ATG3, suggesting the ATG12^A138V^ variant may interrupt ATG3 association with the ATG12–ATG5 complex. In contrast, the p.Phe108 residue lies directly adjacent to ATG5, suggesting the ATG12^F108L^ variant may affect ATG12–ATG5 heterodimer formation, with previous work showing that mutation of the p.Phe108 residue can affect ATG12 stability and conjugation to ATG5 in mouse embryonic fibroblasts.^44^ While the p.Tyr120 of ATG12 is relatively close to ATG5, it is not adjacent to its surface, suggesting a less damaging effect on heterodimer formation. Among over 2000 ATG12 orthologues, Tyr120 is conserved among 44% of sequences, whereas Ala138 is nearly invariant at 89% of sequences.^43^ The Tyr120Ser and Ala138Val substitutions may have an impact on ATG12 interaction with both ATG5 and ATG3. Relevant to this, a substitution at Ala138 (i.e. Ala138Arg) showed a minor impairment of ATG12–ATG5-ATG16L1 binding to ATG3, and abolished LC3 lipidation *in vitro*.^44^

### *ATG12* variants impair autophagy flux and intracellular degradation in patient-derived fibroblasts

Following the results on ATG12 interactions with its binding partners, we performed functional assessment of autophagy by immunoblotting for autophagy-related proteins in control and available *ATG12* subject-derived fibroblasts. Under basal conditions, both S2 and S3 showed stable levels of the core autophagy effectors ATG7 and ATG3, along with stable levels of SQSTM1/p62, an autophagy receptor and substrate (**Fig**. **3A**). S2, however, exhibited a clear loss of the ATG12–ATG5 dimer, along with no detectable ATG12 monomer, likely indicating that decrease in the ATG12–ATG5 conjugate is due to a loss of ATG12. In contrast, cells from S3 revealed stable levels of the ATG12–ATG5 conjugate and ATG12 monomer. Further probing identified the presence of unconjugated ATG5 monomer in S2 only (**Fig. 3A**).

**Figure 3.**
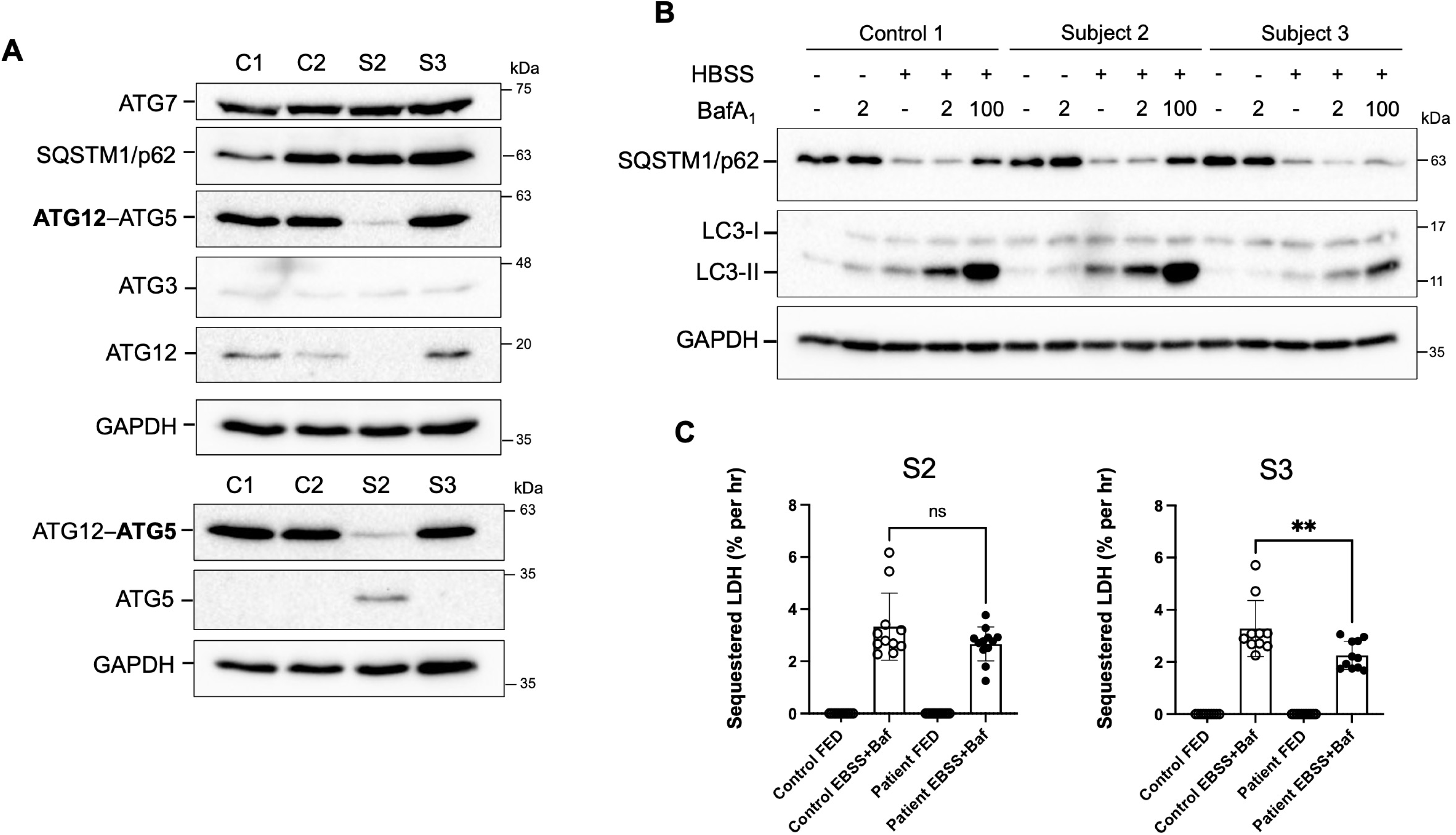
Immunoblot analysis of subject-derived fibroblasts show decreased ATG12 levels or altered autophagy flux. (**A**) Immunoblot of steady state levels of autophagy-related proteins in controls (C1 and C2) and patient-derived fibroblasts (S2 and S3). Immunoblots show decreased ATG12–ATG5 conjugate and loss of unconjugated ATG12 monomer in cells from S2, but not S3. Additional blots show the presence of unconjugated ATG5 in S2, but not in S3. Labels in bold denote the antibody used for detection of the conjugate, (*n*=3) (**B**) Immunoblot of autophagy flux assay. Subject-derived cells were grown under basal conditions or starvation conditions (HBSS) to induce autophagy and treated with 2 nM or 100 nM Bafilomycin A_1_ to block late-stage autophagy for 3 hours before harvesting to probe the rate of LC3-II production. S2 fibroblasts showed minimal change, while S3 fibroblasts showed attenuation of LC3-II production compared to the control fibroblasts. (*n*=3) (**C**) Quantification of LDH sequestration per hour under basal or starvation conditions (EBSS) with lysosome acidification inhibited by 100 nM Bafilomycin A_1_ for 3 hours. Only S3 cells showed a significant decrease in bulk autophagic sequestration. (*n*=3) (\*\**p* < 0.01 Ordinary one-way analysis of variance and Sidak’s multiple comparison test).

Furthermore, we performed a comparative assessment of autophagy flux. Cells were cultured in standard media or under starvation conditions (HBSS) to induce autophagy. We monitored LC3 turnover using a non-saturating concentration of the vacuolar type H^+^-translocating ATPase inhibitor bafilomycin A_1_ for three hours to block autophagic flux at a late stage.^45,46^ Under non-starvation conditions a 2 nM concentration of BafA_1_ resulted in a minimal increase in the lipid-conjugated form of LC3, LC3-II, indicating the dose was non-saturating for both the control and Subject-derived fibroblasts (**Fig. 3B**). Under starvation conditions a 2 nM BafA_1_ treatment resulted in a robust increase in LC3-II in the control and S2 fibroblasts; however, the LC3-II increase was markedly reduced in S3 fibroblasts compared to the control and S2 cells. In addition, a saturating dose of 100 nM BafA_1_ resulted in further accumulation of LC3-II, with S2 fibroblasts being comparable to the control. While 100 nM BafA_1_ increased LC3-II accumulation in S3 fibroblasts, it was still reduced compared to the control and S2. The levels of SQSTM1/p62 did not change in both S2 and S3 fibroblasts and decreased after starvation, suggesting autophagy was suppressed only modestly, even in S3 cells. To complement these findings, we also directly measured non-selective autophagy flux using the classical LDH sequestration assay.^47^ Bulk autophagic sequestration was decreased in cells from S3, whilst flux appeared unaffected in cells from S2 (**Fig**. **3C**).

### ATG12 F108L and A138V variants exhibit modest autophagy defects, while R79Ter and 363+3 A>T variants exhibit profound autophagy defects

To investigate the effects of the subject’s ATG12 variants on autophagy, we conducted rescue experiments. *ATG12* knockout (KO) HeLa cells were generated using the CRISPR-Cas9 system, and successful knockout was confirmed by DNA sequencing (**Supplementary** Fig. 1). Next, the autophagy flux reporter HaloTag–LC3 and each of the 3xHA-ATG12 variants (WT, F108L, Y120S, R79Ter, A138V, and 363+3A>T) were stably expressed (**Fig. 4A**).^30^ We inserted intron 3 into all the *ATG12* cDNAs to evaluate a potential splicing defect of 363+3A>T. Significant amounts of unconjugated ATG12 of the WT and A138V were detected, indicating the overexpression of these constructs after successful splicing. The levels of the ATG12–ATG5 conjugate were similar to the endogenous levels, likely because the amount of ATG5 is rate-limiting. The expression levels of the F108L and Y120S were slightly reduced, while those of R79Ter and 363+3A>T were profoundly reduced (**Fig. 4B, C**). These data suggest that the F108L, Y120S, and R79Ter mutations destabilize ATG12. Furthermore, the minimal expression of the intron variant 363+3A>T confirmed impaired splicing.

**Figure 4.**
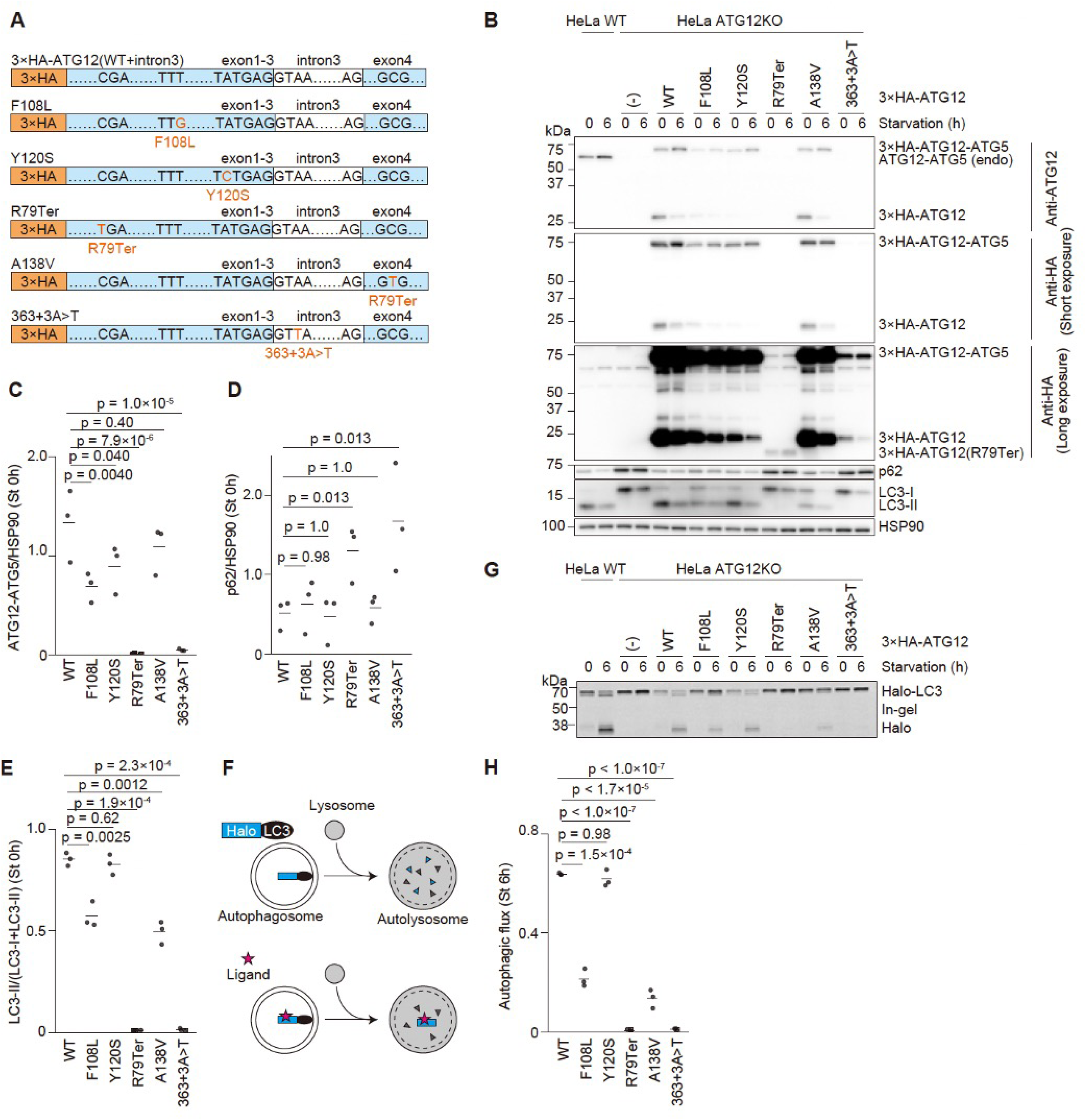
Autophagic activity of the ATG12 variants. (**A**) A structural diagram of the plasmids encoding 3×HA-tagged wild-type human ATG12 and its variants containing intron 3. (**B**) The expression of endogenous ATG12-ATG5, exogenous 3×HA-ATG12 variants, LC3, and p62 before and after 6-h starvation. (**C-E**) Quantification of the levels of ATG12–ATG5 (**C**), p62, (**D**) and LC3 (**E**) shown in (B). Horizontal lines indicate the means, and dots indicate the individual data from three independent experiments. Data were statistically analysed using one-way ANOVA with Tukey test. (**F**) Schematic representation of the HaloTag–LC3 processing assay. HaloTag, which is normally degraded in lysosomes, becomes stable after ligand binding. (**G**) In-gel fluorescence detection of the HaloTag fragment in HaloTag-LC3-expressing HeLa cells pulse-labeled with 100 nM TMR-conjugated ligand for 20 minutes. Cells were starved for 6 h. (**H**) Quantification of the results shown in (G). HaloTag^TMR^ band intensity was normalized by the sum of the band intensities (HaloTag^TMR^-LC3 and HaloTag^TMR^). Horizontal lines indicate the means, and dots indicate the individual data from three independent experiments. Data were statistically analysed using One-way ANOVA with Tukey test.

Next, we evaluated the autophagic flux of these rescued cells.^48^ SQSTM1/p62, a well-established selective receptor and substrate of autophagy, accumulated in *ATG12* KO cells, but not in cells rescued with wild-type ATG12.^49^ The expression of SQSTM1/p62 was also returned to basal levels in cells expressing F108L, Y120S, or A138V, but remained elevated in cells expressing R79Ter or 363+3A>T (**Fig. 4B, D**). LC3 lipidation was rescued in cells expressing Y120S, whereas it was partially impaired in cells expressing F108L or A138V and was not detected in cells expressing R79Ter or 363+3A>T (**Fig. 4B, E**). To quantify autophagic flux more precisely, we performed the HaloTag–LC3 processing assay.^30^ When HaloTag–LC3 is delivered to lysosomes via autophagy, it is efficiently degraded. However, after binding to its ligands, such as the tetramethylrhodamine (TMR)-conjugated ligand, HaloTag becomes resistant to lysosomal degradation, allowing the HaloTag fragment to accumulate depending on autophagic activity (**Fig. 4F)**. The HaloTag fragment was not detected in *ATG12* KO cells but was detected in cells expressing wild-type ATG12 after 6-h starvation (**Fig. 4G, H**). The amount of the HaloTag fragment decreased in cells expressing F108L or A138V and was virtually undetectable in cells expressing R79Ter or 363+3A>T. These results suggest that F108L and A138V exhibit significantly reduced autophagic activity, and R79Ter and 363+3A>T exhibit virtually no autophagic activity.

In contrast, cells expressing Y120S demonstrated normal HaloTag processing (**Fig. 4G, H**), suggesting intact autophagic activity in this experimental setting. However, given that the expression level of Y120S decreased in patient-derived fibroblasts (obtained from S2), Y120S may have an autophagy defect at the endogenous expression level, which could be overcome by its overexpression.

### F154L, W166S and A184V mutations of Atg12 impaired Atg8 lipidation and autophagy flux under acute nitrogen starvation in yeast

Our analyses of autophagic flux in subject-derived primary cells revealed subtle biochemical deficits. As *ATG12* and the autophagy process are evolutionarily conserved, we were able to use complementation studies in *S. cerevisiae* expressing homologous *Atg12* variants to assess their effects on autophagy *in vivo*. Sequence alignments demonstrated ATG12^F108^, ATG12^Y120^ and ATG12^A138^ of the human ATG12 correspond to ATG12^F154^, ATG12^W166^ and ATG12^A184^ of yeast Atg12 (**Fig**. **5A**). We constructed *atg12*Δ yeast cells and complemented them with 3xHA-Atg12, 3xHA-Atg12^F154L^, 3xHA-Atg12^W166S^, or 3xHA-Atg12^A184V^, via integrative plasmids. As Atg12 can conjugate with Atg5 and function as an E3-like enzyme to catalyse the lipidation reaction of Atg8, we first examined if starvation-induced Atg8 lipidation was affected by these mutations. The attachment of the lipid molecule phosphatidylethanolamine (PE) at the C-terminal end of Atg8 results in faster electrophoretic mobility in urea-SDS-PAGE gels relative to the nonlipidated form. No Atg8 lipidation occurred in the empty vector (–) transformant, whereas robust Atg8 lipidation was observed in the *atg12Δ* cells transformed with the 3xHA-Atg12 (WT) plasmid (**Fig. 5B**). The 3xHA-Atg12^A184V^ mutant partially rescued Atg8 lipidation defects, but to a lesser extent than 3xHA-Atg12. The 3xHA-Atg12^F154L^ and the 3xHA-Atg12^W166S^ mutants caused more severe defects in Atg8 lipidation.

**Figure 5.**
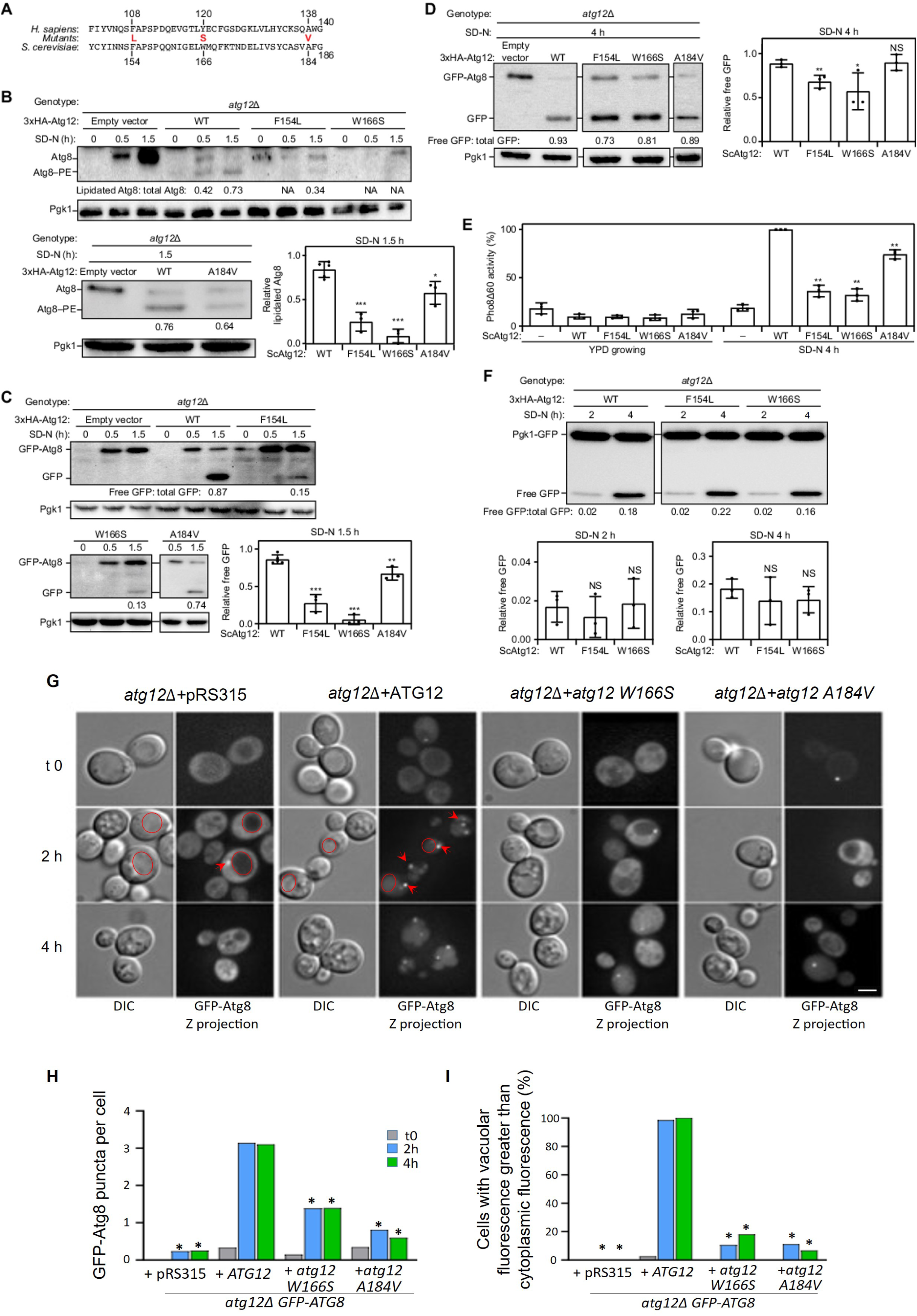
Disease-associated mutations of Atg12 impaired Atg8 lipidation and autophagy flux under acute nitrogen starvation in yeast. (**A**) Alignment of the human (UniProtKB: O98417) and yeast ATG12 (UniProtKB: P38316) sequences covering the mutated amino acids F154L, W166S and A184V. (**B**) WLY176 *atg12*Δ cells transformed with *pRS406* empty vector, *pRS406-ATG12p-3xHA-ATG12*, *pRS406-ATG12p-3xHA-ATG12[F154L]*, or *pRS406-ATG12p-3xHA-ATG12[W116S]* were grown to mid-log phase in YPD (0 h), centrifuged and resuspended in SD-N medium for the indicated times. Cell lysates were prepared, subjected to 18% urea-SDS-PAGE, and analysed by western blot. The ratio of lipidated Atg8 to total Atg8 (lipidated Atg8 plus nonlipidated Atg8) was quantified to indicate the Atg8 lipidation level. (**C-D**) Cell lysates were prepared, subjected to 10% SDS-PAGE, and analysed by western blot. The ratio of free GFP to total GFP (free GFP plus GFP-Atg8) was quantified to indicate autophagy flux. Pgk1 was used as a loading control. (**E**) Cell lysates were prepared and analysed by the Pho8Δ60 assay. Pho8Δ60 activity was normalized to WLY176 *atg12*Δ cells transformed with *pRS406-ATG12p-3xHA-ATG12* in SD-N for 4 h (set to 100%). (**F**) SEY6210 *atg12*Δ cells transformed with *pRS406* empty vector, *pRS406-ATG12p-3xHA-ATG12*, *pRS406-ATG12p-3xHA-ATG12[F154L]*, or *pRS406-ATG12p-3xHA-ATG12[W116S]* were grown to mid-log phase in YPD (0 h), centrifuged and resuspended in SD-N medium for the indicated times. Cell lysates were prepared, subjected to 10% SDS-PAGE, and analysed by western blot. The ratio of free GFP to total GFP (free GFP plus Pgk1-GFP) was quantified to indicate autophagic cargo degradation. In this figure, summary data were presented as the mean ± SD. \**p* < 0.05, \*\**p* < 0.01, \*\*\**p* < 0.001, and NS, not significant; Student’s *t*-test. For all western blot and Pho8Δ60 experiments, (n=3-5). (**G**) GFP-Atg8 live imaging in an *atg12*Δ strain transformed with plasmids expressing Atg12 or mutated Atg12. Cells were collected before (t0), and at 2 and 4 h after nitrogen starvation. Red arrows indicate autophagosomes or phagophores and red circles indicate the yeast vacuole (scale bar: 3 µm). Graphs show average dots per cell and the percentage of cells with increased vacuolar GFP signal. Significant average dots per cell differences relative to the *atg12*Δ strain expressing the ATG12 plasmid (\**p* < 0.001 Kruskall-Wallis test) (*n*=3). Significant vacuolar fluorescence with the *atg12Δ* strain expressing the ATG12 plasmid (*n*=3).

To quantitatively characterize the effects of these mutations on bulk autophagy flux, we performed the GFP-Atg8 processing assay: in brief, a portion of GFP-tagged Atg8 bound to the inner side of the autophagosome is delivered to the vacuole via autophagy. The GFP is relatively resistant to vacuolar hydrolytic enzymes in comparison to Atg8, and the free GFP can be resolved and detected by western blot. The ratio between free GFP versus total GFP reflects the flux of vacuolar delivery of the autophagosomes. Under acute nitrogen starvation (SD-N 0.5 h and SD-N 1.5 h), there was no GFP-Atg8 processing signal in the empty vector transformants, but the free GFP cleavage from the GFP-Atg8 fusion protein was abundant in the *atg12*Δ cells transformed with the WT 3xHA-Atg12 plasmid (**Fig. 5C**). In contrast, the 3xHA-Atg12^F154L^, or 3xHA-Atg12^W166S^ transformants only partially rescued the GFP-Atg8 processing defect in the *atg12*Δ cells.

We observed that during prolonged nitrogen starvation (SD-N, 4 h), the negative effects of the mutations became milder (**Fig. 5D**). We reasoned that these mutations only partially affect the functions of the conjugating enzyme. As the starvation time increases, the total autophagy flux becomes similar to that of the WT, which has reached a plateau.

In the second autophagy flux assay, Pho8Δ60 is an engineered form of the Pho8 phosphatase whose vacuolar delivery is autophagy dependent. Once entering vacuoles, the Pho8Δ60 propeptide is cleaved and activated. Pho8Δ60 activity was low under nutrient-rich conditions among all three transformants (**Fig. 5E**). After 4 h in the nitrogen-starvation medium, robust Pho8Δ60 activity was induced in the 3xHA-Atg12 transformants, but not the empty vector transformants. The Pho8Δ60 activity was lower in all three mutants, supporting the hypothesis that these mutations in Atg12 impaired autophagy flux.

We also performed a Pgk1-GFP processing assay to further examine the effects of the mutations on autophagic cargo degradation. However, these mutations did not affect Pgk1-GFP processing (**Fig. 5F**), likely because its degradation primarily occurs during prolonged starvation, when the autophagy flux defect caused by the mutations is less pronounced.

Autophagic activity can also be quantitatively measured using microscopy. GFP-Atg8 delivery to the vacuole via autophagosomes results in GFP puncta accumulating inside the vacuole, indicating autophagic activity. Punctate GFP signals indicative of autophagosomal structures were still able to form in strains harbouring Atg12^W166S^ and Atg12^A184V^ variants; however, their number was greatly decreased (**Fig**. **5G, H**). Additional quantification of GFP presence in the vacuole also confirmed a reduction in the delivery of GFP to the degradative compartment (**Fig**. **5I**).

### Loss of *atg12* function in zebrafish causes developmental delay, impaired brain function, and pre-adult lethality

To investigate the function of ATG12 in vivo, we generated a knockout in the zebrafish ortholog, *atg12,* using CRISPR/Cas9-mediated mutagenesis. A 10-bp deletion within exon 2 of the *atg12* gene was identified and confirmed by Sanger sequencing (**Fig. 6A**). This frameshift mutation is predicted to truncate the ATG12 domain, resulting in a loss-of-function allele. Morphological analyses revealed that *atg12^-/-^* larvae were morphologically indistinguishable from their *atg12^+/+^* siblings at 5 days post-fertilization (dpf). However, as development progressed, *atg12^-/-^* larvae exhibited developmental delay, and by 15 dpf, they were notably smaller than their wild-type siblings (**Fig. 6B**). Genotyping analyses showed that *atg12^-/-^* larvae were present at Mendelian ratios at 5 dpf but progressively declined by 60 dpf, indicating that homozygous mutants fail to survive to adulthood (**Fig. 6C**). These findings suggest that *atg12* is essential for post-larval growth and viability in zebrafish.

**Figure 6.**
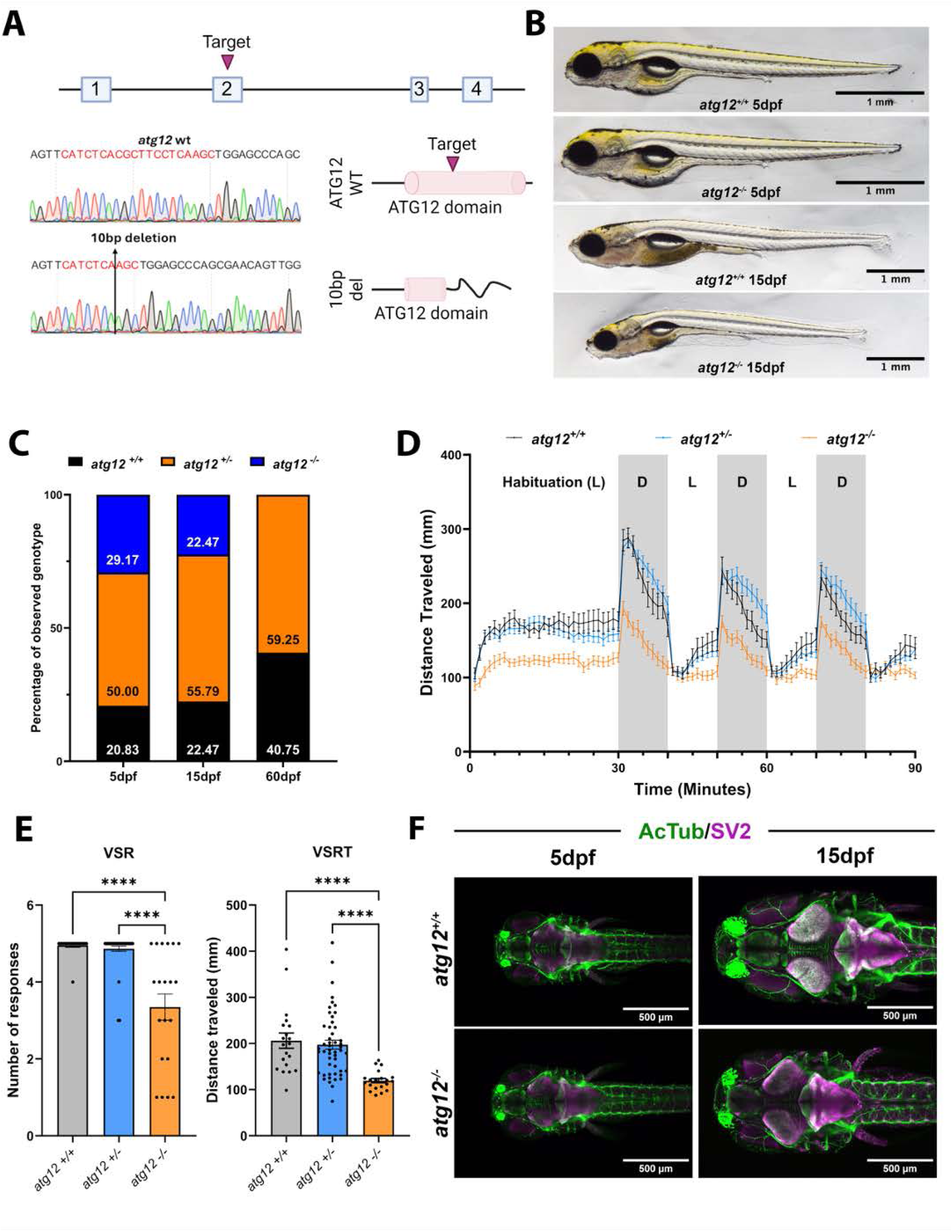
Loss of *atg12* impairs zebrafish development, brain function, and survival. (**A**) CRISPR/Cas9-mediated mutagenesis generated a 10-bp deletion within exon 2 of *atg12*, confirmed by Sanger sequencing. The resulting frameshift is predicted to truncate the ATG12 domain, producing a loss-of-function allele. (**B**) Bright-field images of *atg12+/+* and *atg12*-/-larvae at 5 and 15 dpf. Mutants are morphologically indistinguishable from wild-type at 5 dpf but show developmental delay and reduced body size by 15 dpf. Scale bars, 1 mm. (**C**) Genotypic distribution of *atg12+/+*, *atg12+/-*, and *atg12*-/-larvae at 5, 15, and 60 days post-fertilization (dpf). Homozygous mutants were present at Mendelian ratios at 5 dpf but progressively declined by 60 dpf, indicating decreased post-larval survival. (**D**) Locomotor activity during alternating light (L) and dark (D) phases. Wild-type larvae exhibited robust increases in movement following light-to-dark transitions and subsequent habituation, whereas *atg12*-/-mutants showed significantly reduced activity throughout both phases. (**E**) Quantification of visual startle responses (VSR). Both the number of responses (VSR) and total distance traveled (VSRT) were significantly reduced in *atg12*⁻/⁻ larvae compared with wild-type and heterozygous siblings. Data represent mean ± SEM. ****p < 0.0001 by one-way ANOVA with post hoc comparisons. (**F**) Light-sheet images of acetylated tubulin (AcTub, green) and synaptic vesicle protein 2 (SV2, magenta) in *atg12*⁺/⁺ and *atg12*⁻/⁻ larvae at 5 and 15 dpf. Mutants exhibit reduced brain size and reduced synaptic intensity at 15 dpf. Scale bars, 500 μm.

To assess the impact of *atg12* deficiency on neurodevelopment in zebrafish, we evaluated behavior assays to measure locomotor responses in the light-dark cycle (LDC) and visual startle response (VSR) assays. In the LDC assay, wild-type larvae displayed robust locomotor activity characterized by increased distance travelled during dark phases and subsequent habituation across cycles (**Fig. 6D**). In contrast, *atg12^-/-^* mutants exhibited markedly reduced locomotor activity throughout both light and dark phases. Consistent with these findings, *atg12^-/-^* larvae demonstrated significantly attenuated visual startle responses, with both the number of responses (VSR) and total distance travelled (VSRT) significantly reduced compared to wild-type controls (**Fig. 6E**). Together, these results indicate that loss of Atg12 impairs sensorimotor function and arousal responses, leading to reduced locomotor output and impaired responses to changes in visual stimuli.

To further examine the mechanisms behind the observed functional deficits in *atg12^-/-^* larvae, we analysed the 5 dpf and 15 dpf larval brain morphology using immunohistochemistry. Larvae were stained for acetylated tubulin (AcTub) and the synaptic vesicle marker SV2. At 5 dpf, *atg12^-/-^*larvae showed no apparent differences in brain morphology or axonal organization compared to wild-type siblings. However, by 15 dpf, mutants exhibited a smaller overall brain size (**Fig. 6F**), in line with the global development delay observed in affected individuals.

In summary, our data demonstrate that *Atg12* is required for normal growth, brain development, and neural function in zebrafish. These findings provide functional evidence linking *Atg12* disruption to impaired neurodevelopment, supporting a potential role for *Atg12* dysfunction in the pathophysiology of human neurodevelopmental disorders.

## Discussion

Here we report six patients from five unrelated families harbouring variants in the core autophagy gene *ATG12*, a gene not previously associated with a human phenotype. The patients share clinical features of ataxia and global developmental delay with the neuroradiological finding of profound cerebellar hypoplasia. For comparison with the phenotypes of previously reported patients with *ATG5* and *ATG7* pathogenic variants (**Supplementary Table 5**).

Our *in-silico* analyses using published x-ray crystal structures, combined with our AlphaFold structural modelling, suggest that the p.Tyr120Ser and p.Ala138Val variants map to a ubiquitin-like fold in ATG12, with the p.Phe108Leu variant being previously shown to affect ATG12 conjugation and function.^44^ While we observed loss of the stable ATG12–ATG5 complex in S2 cells, this was not observed in S3 cells, although only S3-derived fibroblasts showed reduced autophagic flux. Additionally, *ATG12* KO HeLa cells and *atg12* null yeast cells are unable to fully recover autophagy when complemented with equivalent missense ATG12 (except Y120S) and *Atg12* variants, respectively, suggesting the identified variants have a negative effect *in vivo.* Regardless, while the human *ATG12* and yeast *Atg12* variants retain partial function, the mild autophagy attenuation observed across the multiple functional assays strongly suggests that these variants are detrimental to autophagy. This finding, combined with the neurological phenotypes observed in both families, suggests that the autophagy impairment stemming from *ATG12* variants is incompatible with sustaining neural integrity in humans. Zebrafish functional studies identify a critical role for Atg12 in zebrafish development and neural function. Disruption of *atg12* through CRISPR/Cas9-mediated mutagenesis resulted in a loss-of-function allele that caused profound post-larval growth delay, reduced locomotor activity, and pre-adulthood lethality. Despite normal early morphology at 5 days post-fertilization (dpf), *atg12^-/-^* larvae exhibited progressive defects in neuronal organization, reduced synaptic labelling, and impaired sensorimotor responses. In humans, deficits in these processes are characteristic of neurodevelopmental disorders, including ataxia, intellectual disability, and epilepsy, where alterations in sensory responsiveness, motor coordination, and habituation are commonly observed. Thus, the zebrafish *atg12* mutants provide a tractable in vivo model to investigate how defective autophagy contributes to these phenotypes.

Decades of work across various model systems have elaborated the intricate molecular regulation and function of canonical macroautophagy. As part of this work, numerous knockout mouse models of various *ATG* genes have been produced and characterised, including models of *atg12*, *atg5, atg3,* and *atg7*.^13–15,50^ Global knockout models of these genes involved in the ATG conjugation system result in perinatal lethality, inferring that deleterious variants in these genes are incompatible with life. However, in addition to these results involving *ATG12,* recent publications from our group and others have described cases of patients harbouring pathogenic variants in *ATG5* and *ATG7* surviving past birth, with some patients reaching ages towards the population’s life expectancy.^8–10^ This observation suggests a potential for supplemental or compensatory pathways in humans that may not be present in shorter-lived model organisms such as mice. It is important to remember that previous work has suggested autophagy is generally essential in mammals; mutant alleles resulting in severe defects may therefore result in non-viable embryos, favouring the detection of mutations that result in more subtle phenotypes.^51^ Nevertheless, the neuropathology observed in a preclinical CNS-specific *atg7* conditional knockout model, together with those reported here in human *ATG12* patients, underscores the essential role of autophagy in maintaining neural integrity in mammals.^9–11^

An additional feature of interest in these *ATG12* cases is the consistency in neuroradiological findings with the reported *ATG7* and *ATG5* cases (**Supplementary Table 5**).^8,9^ The hypoplasia of the corpus callosum observed in S1, S2, S4 and S5 is also consistent with the *ATG7* and *ATG5* cohorts, as is the cerebellar hypoplasia observed. ^8,9,52^ It is unclear as to why these areas appear to be most susceptible to damage, but the consistency across the cohort would suggest these areas are more reliant on autophagy during development. More specifically these regions, especially the cerebellum, may be more reliant on mitophagy, a form of selective autophagy specific to mitochondria, during development. Previous work using the *mito*-QC reporter mouse has demonstrated that cerebellar Purkinje cells exhibit high levels of mitophagy in the cerebellum of mice.^53^ Additionally it has been demonstrated that mitophagy rates in cerebellar Purkinje cells and external granule cells of ageing mice increase; however, *auto*-QC analysis showed that bulk autophagy does not follow the same trend.^54^ This finding demonstrates the disparity between autophagy and mitophagy in cerebellar cell types and may hint that certain forms of selective autophagy are the predominant drivers for the specificity of the phenotypes associated with congenital *ATG* disorders. Similar work investigating the selective autophagy rates in contrast to bulk autophagy in the developing brain may help to elucidate this.

Given the similarities between the various *ATG* cohorts (**Supplementary Table 5**), it would be interesting to ascertain if a generalised autophagy modulation therapy would be effective for all three of the described disorders, or whether they would require more specific interventions, such as gene therapy approaches. However, given the clear developmental component in these infantile-onset disorders, further insights are needed to identify intervention points that could offer maximum benefit.

The range of phenotype severity in the cohort is also interesting. Fibroblasts from S2 were found to have LC3 lipidation comparable to that of the control in autophagy flux assays, despite their loss of the ATG12–ATG5 complex, likely due to the minimal amount of the ATG12– ATG5 conjugate required for efficient lipidation of ATG8 proteins.^55^ However, the corresponding subjects were unable to survive past early childhood, with S1 dying of intractable infantile spasms and epileptic encephalopathy before the age of 1 year, and sibling S2 also being diagnosed with epileptic encephalopathy and dying in early childhood. In contrast, S3 was found to maintain their ATG12–ATG5 conjugate, but S3-derived fibroblasts had a severe reduction in LC3-I lipidation when analysed by an autophagy flux assay. Despite this, S3 appears to have a less severe neurological phenotype, but with the complication of kidney disease and other non-neurological phenotypes. The kidney disease phenotype was also observed in his sibling, not available for genetic testing, suggesting he too may have been affected with the same genetic variants. The seemingly paradoxical relationship between biochemical defect and patient phenotype is not without precedent. Of the previously described *ATG7* patient cohort, S1 and S2 (segregating loss of function variants) had the most severe biochemical defect with undetectable levels of ATG7 and no detectable LC3-II production, yet presented with the mildest phenotype.^8^ In contrast, S3 and S4 (segregating missense variants) had detectable levels of ATG7 and LC3-II production by immunoblot, yet presented with a more severe neurological phenotype.^8^

In Family 2, both S3 and an untested sibling presented with kidney disease. This phenotype could represent a less common manifestation of ATG12 dysfunction or may have an unrelated genetic aetiology, especially considering parental consanguinity in this family, which increases the risk for recessive disorders. Despite extensive analysis, WES did not reveal any putative pathogenic variants that could explain the kidney phenotype.

It is possible that the early lethality observed in Family 1 may have prevented a kidney disease phenotype from developing, however its absence from other members of this and the *ATG7* and *ATG5* cohorts would appear to make this less likely. It is hoped that this report of *ATG12*-related pathology will result in *ATG12* featuring in prospective diagnostic testing algorithms and further expansion of the genotype-phenotype correlation may help elucidate the potential renal involvement as additional families are diagnosed and reported. Similarly, for early lethality, although S1 and S2 had a rapidly progressive and fatal disease course, only one other *ATG* patient is reported to have the same severe phenotype, from the *ATG7* cohort, whilst other *ATG* patients have a milder disease course.^8^ It is likely that genetic diagnoses of *ATG* disorders will increase with the widespread use of next-generation sequencing in the diagnostic process.

Another phenotypic similarity to the *ATG7* cohort is optic atrophy, found in 7/9 investigated patients, as in patient S3.^8^ Nystagmus in patients S2 and the deceased sibling of S3 may have resulted from optic atrophy, although this remains uncertain. Sensorineural deafness is also reported in 2/12 patients in the *ATG7* cohort, as in patient S3 (**Supplementary Table 5**).^8^ Together, this demonstrates that while the main overlapping phenotypes remain the cerebellar hypoplasia and ataxia, there are other similarities between *ATG*-related disorders, which will no doubt expand with the identification of new patients.

The phenotypes of patients with pathogenic variants in the *ATG5*, *ATG7,* and *ATG12* genes are clearly distinct from the phenotypes of BPAN (beta-propeller protein-associated neurodegeneration) patients with mutations in *WDR45* (*WIPI4*).^56^ *WDR45* (*WIPI4*) is one of the four human homologs of yeast ATG18, and it functions with ATG2. BPAN is characterized by iron deposition in the globus pallidus and the substantia nigra and presents with non-progressive psychomotor retardation during childhood and rapidly progressing Parkinson-like symptoms in adulthood. Since these symptoms differ significantly from those of diseases caused by pathogenic variants in genes related to the ATG conjugation system, it is possible that the impairment of non-autophagy functions of WDR45/WIPI4 contributes to the onset of BPAN.^57^

Next generation sequencing efforts have been instrumental in revealing novel congenital autophagy-associated defects that were once thought to be incompatible with life.^58^ Although still exceedingly rare, our findings extend the repertoire of genetic variants linked to autophagy dysfunction and further emphasize the clinical significance of autophagy for human nervous system health and disease. We anticipate that the discovery and profiling of additional *ATG* variants will pave the way for therapeutic interventions aimed at ameliorating disorders associated with autophagic dysfunction.

## Data availability

WES data are available upon reasonable request. Distribution of sensitive data is subject to specific restrictions.

## Data Availability

WES data are available upon reasonable request. Distribution of sensitive data is subject to
specific restrictions.

## Acknowledgements

We are grateful to the families for participating to the study. We thank Dr. Siren Berland and co-workers at Haukeland Universitetssjukehus, Bergen, Norway, for performing genetic testing in a clinical setting. We thank the staff of the HiLIFE electron microscopy unit (directed by Dr. Eija Jokitalo) for their excellent technical assistance, and Dr. Helena Vihinen for insightful advice on sample preparation. We are grateful to Professor Eeva-Liisa Eskilinen (University of Turku, FI) for insightful discussions and guidance on discriminating autophagic sub-compartments. We thank Yongheng Liang (Nanjing Agricultural University) for plasmid *pP1kGFP-ATG8(406)* and Florence Piron Prunier for technical advice. We also thank UNINETT Sigma2, the National Infrastructure for High Performance Computing and Data Storage in Norway, for support to store and analyse High-Throughput Sequencing data.

## Author contributions

JL, SA, YH, FS, TE and MP carried out the experimental assessments of autophagy in patient-derived cells and model systems. JL and TJM performed structural modelling studies. AI, EH, DM, GR, PV, RM, HH and CLA performed the molecular genetic analyses. CP, KH and GKV performed functional studies in zebrafish. DW and KF undertook and assessed the neuroradiological assessments. HM, AB, MFS, CK, EF, SN, AS, RS, FR, SM, VU, RMcF supported the clinical investigations of all families. JL, JJC, RM, WWY, DJK, RL, NM, TGM, MO, CLA and RWT interpreted all the data. RWT, WWY, DJK, RL, NM, TGM, MO and CLA supervised the study and acquired funding. JL, SA, YH, DJK, NM, GKV, TGM, MO, CLA and RWT wrote the manuscript. All authors contributed to the revision of this manuscript draft.

---

**Figure S1.**
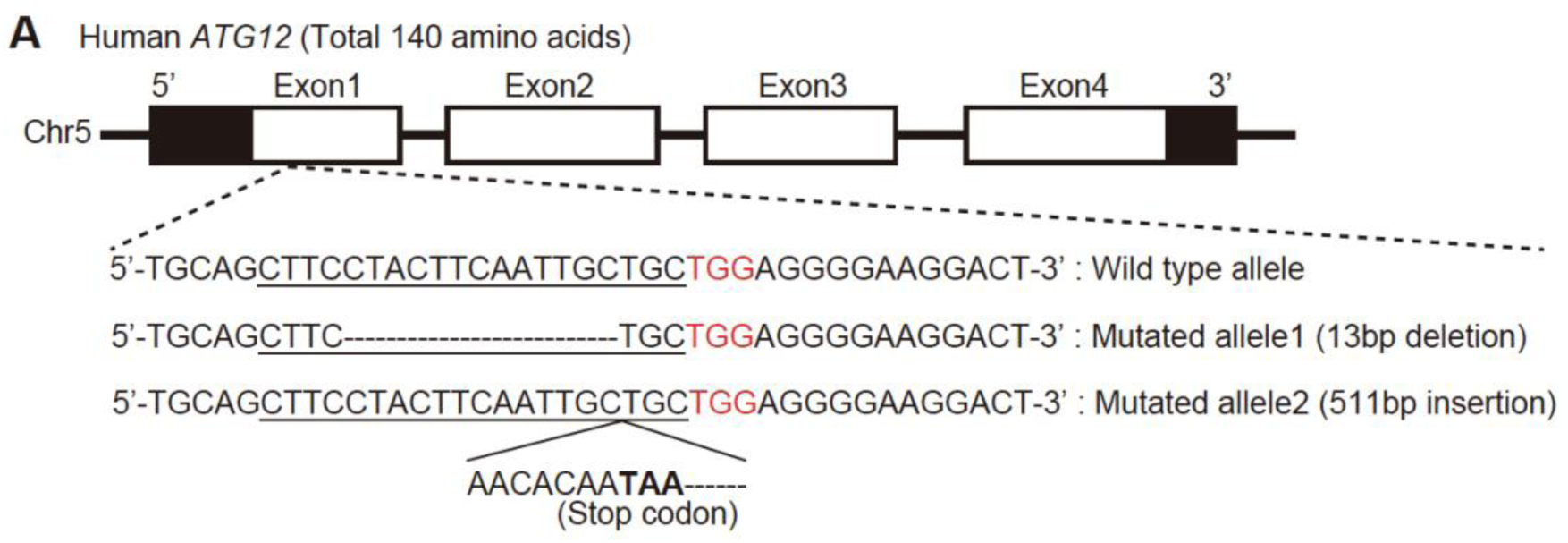
Generation of *ATG12* knockout cells. (**A**) Schematic representation of the Cas9-gRNA-target site (underlined) in the *ATG12* genomic locus. The protospacer-adjacent motif sequence is shown in red. A deletion of 13 bp and an insertion of 511 bp in the knockout allele are indicated.

**Table S1.**
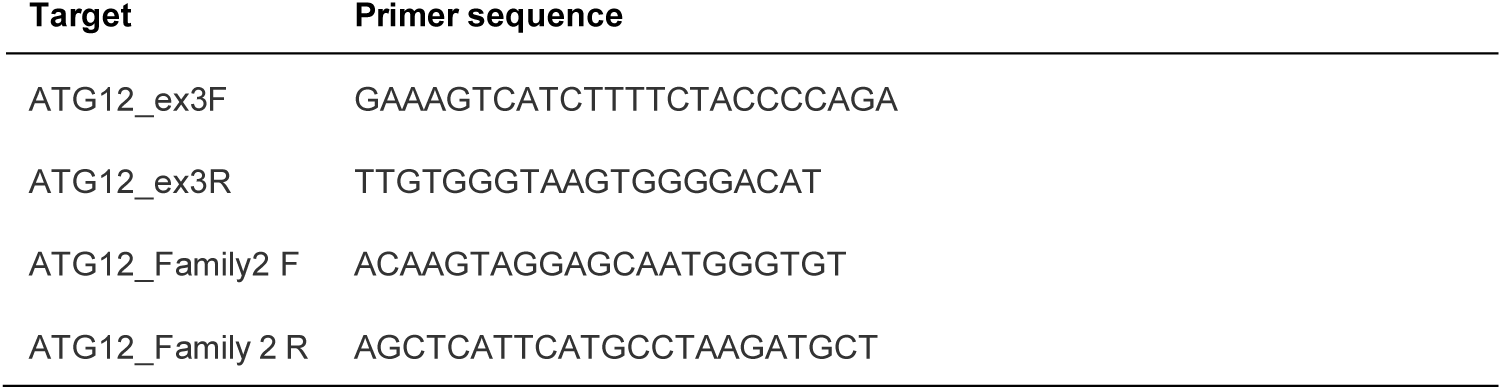
Primers used for genomic DNA confirmation of patient variants.

**Table S2.**
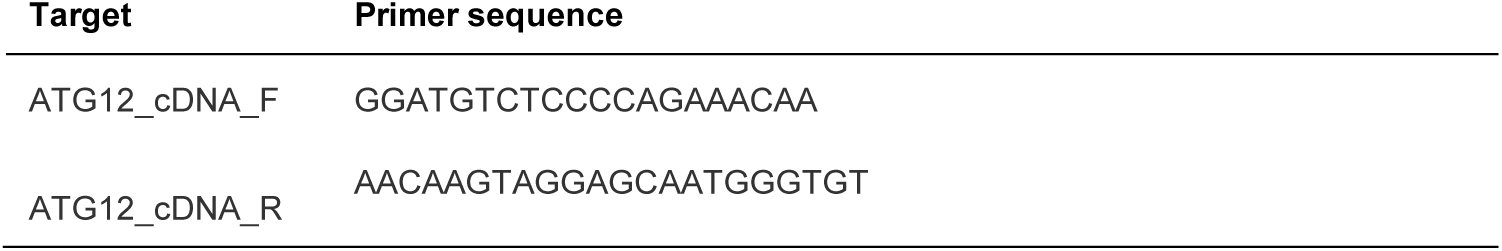
Primers used for cDNA investigations into splice variant.

**Table S3.** Genotype and source of *S. cerevisiae* used in this study.

| Strain | Genotype | Source | Purpose |
| --- | --- | --- | --- |
| JH914 | SEY6210+*atg12Δ::KANMX6 PGK1-GFP::natMX6* | This study | Pgk1-GFP assay |
| JH947 | WLY176 *pRS405-ATG8p-GFP-ATG8::LEU2 atg12Δ::HIS3MX6 pRS406-ATG12p-3xHA-ATG12 F154L::URA3* | This study | Atg8 lipidation, GFP-Atg8, and Pho8Δ60 assays |
| JH948 | WLY176 *pRS405-ATG8p-GFP-ATG8::LEU2 atg12Δ::HIS3MX6 pRS406-ATG12p-3xHA-ATG12 W166S::URA3* | This study | Atg8 lipidation, GFP-Atg8, and Pho8Δ60 assays |
| JH949 | SEY6210+*atg12Δ::KANMX6 PGK1-GFP::natMX6 pRS406::URA3* | This study | Pgk1-GFP assay |
| JH950 | SEY6210+*atg12Δ::KANMX6 PGK1-GFP::natMX6 pRS406-ATG12p-3xHA-ATG12::URA3* | This study | Pgk1-GFP assay |
| JH952 | SEY6210+*atg12Δ::KANMX6 PGK1-GFP::natMX6 pRS406-ATG12p-3xHA-ATG12 F154L::URA3* | This study | Pgk1-GFP assay |
| JH953 | SEY6210+*atg12Δ::KANMX6 PGK1-GFP::natMX6 pRS406-ATG12p-3xHA-ATG12 W166S::URA3* | This study | Pgk1-GFP assay |
| JH954 | WLY176 *pRS405-ATG8p-GFP-ATG8::LEU2 atg12Δ::HIS3MX6 pRS406-ATG12p-3xHA-ATG12 A184V::URA3* | This study | Atg8 lipidation, GFP-Atg8, and Pho8Δ60 assays |
| JH955 | SEY6210+*atg12Δ::KANMX6 PGK1-GFP::natMX6 pRS406-ATG12p-3xHA-ATG12 A184V::URA3* | This study | Pgk1-GFP assay |
| OC751 | BY4742, *atg12::KanMX4, ura3Δ0, his3Δ1, leu2Δ0, lys2Δ0* | YKO collection | GFP-Atg8 *in-vivo* microscopy |
| OC588 | BY4742, *atg8::GFP-ATG8-URA3, ura3Δ0, his3Δ1, leu2Δ0, lys2Δ0* | This study | GFP-Atg8 *in-vivo* microscopy |
| OC744 | BY4742, *atg8::GFP-ATG8-URA3, atg12::KanMX4, ura3Δ0, his3Δ1, leu2Δ0, lys2Δ0* | This study | GFP-Atg8 *in-vivo* microscopy |
| YLY290 | WLY176 *pRS405-ATG8p-GFP-ATG8::LEU2 atg12Δ::HIS3MX6 pRS406::URA3* | This study | Atg8 lipidation, GFP-Atg8, and Pho8Δ60 assays |
| YLY291 | WLY176 *pRS405-ATG8p-GFP-ATG8::LEU2 atg12Δ::HIS3MX6 pRS406-ATG12p-3xHA-ATG12::URA3* | This study | Atg8 lipidation, GFP-Atg8, and Pho8Δ60 assays |

**Table S4.**
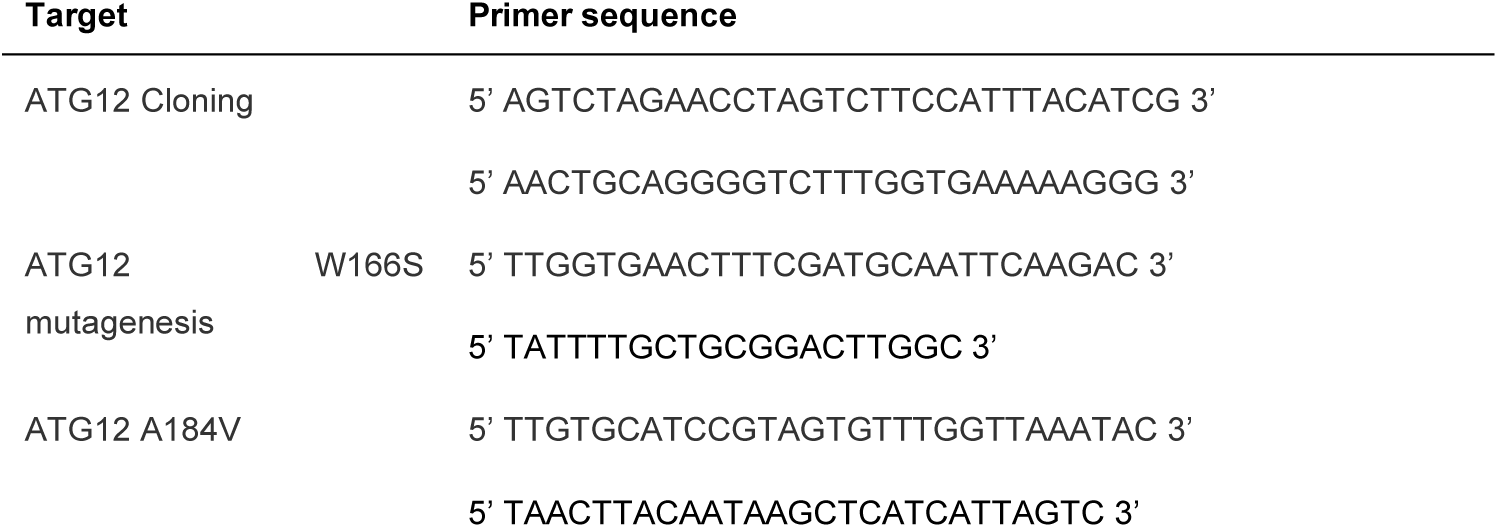
Oligonucleotides used to generate plasmid constructs for yeast complementation studies.

**Table S5.** Overview of phenotypes associated with described congenital disorders in *ATG* genes.

| Gene | *ATG12* |  |  |  |  |  | *ATG7* | *ATG5* |
| --- | --- | --- | --- | --- | --- | --- | --- | --- |
| No. of patients | *n* = 6 |  |  |  |  |  | *n* = 12 | *n* = 2 |
| Reference | Lambton *et al.* present study |  |  |  |  |  | Collier *et al* 2021<sup>8</sup> | Yapici *et al* 2005<sup>10</sup><br>Kim *et al* 2016<sup>9</sup> |
| Subject | S1 | S2 | S3 | S4 | S5 | S6 | S1-12 | Patient 1 and 2<sup>a</sup> |
| Ataxia | NA | NA | + walks with support | Nonambulant | NA | Did not achieve walking | 9/11 | 2/2 |
| DD | + | + | + | + | + | + | 12/12 | 2/2 |
| Intellectual disability | + EE | + EE | + | + | + EE | + | 12/12 | 2/2 |
| Seizures | + IS | + | + | + IS | + | + | 2/12 | 0/2 |
| CC hypoplasia | + | + | - | + | +(Body of cc thin and absent posteriorly) | N/A | 7/9 | 0/2 |
| Cerebellar (vermian) hypoplasia | + | + | + | + | + | N/A | 7/11 | 2/2 |
| Other brain anomaly | Reduced white and grey matter | Deep sulcation occipital lobe | Polymicrogyria (widespread) | Microcephaly with brachycephaly & cerebellocerebral atrophy, thinning of pons, brainstem. hypomyelination | Prominent cavum septum pellucidum, small cerebellar hemispheres, small posterior fossa | N/A | Brain volume loss 3/9 | - |
| Sensorineural deafness | NA | NA | + | NA | NA | N/A | 2/12 | NA |
| Optic atrophy | NA | - | + | + | NA | No but vernal keratoconjunctivitis with bilateral pseudogerontoxon | 7/9 | NA |
| Kidney disease | NA | NA | ESKD | NA | - | - | NA | NA |
| Dysmorphology | NA | NA | + | + | + | - | 4/12 | NA |
| Growth retardation | NA | NA | + | + | + | + | 3/8 | NA |
| Other |  |  | Dry scaly skin, sparse hair, pancreatitis, cholecystitis, inguinal hernia, hypothyreosis |  | Movement disorder with dystonia and dyskinesia, cerebral irritability, PEG fed | Repeated chest and GI infections, failure to thrive, microcephaly |  | Club feet 1/2 |

+ = present; - = absent; CC = corpus callosum; CI = cognitive impairment; DD= developmental delay; EE = epileptic encephalopathy; ESKD = end-stage kidney disease; IS = infantile spasms; m = months, NA = not assessed; S = subject; y = years; PEG = percutaneous endoscopic gastrostomy. <sup>a</sup> Patient 1 and Patient 2 from family 1 in Yapici *et al* 2005; reassessed as subjects 601 and 602 in Kim *et al* 2016

